# Cost-effectiveness of addressing constraints in childhood acute malnutrition management in Malawi using the *Thanzi La Onse* health system simulation framework

**DOI:** 10.64898/2026.03.05.26347696

**Authors:** Eva Janoušková, Ines Li Lin, Emmanuel Mnjowe, Watipaso Mulwafu, Emilia Connolly, Sakshi Mohan, Dominic Nkhoma, Andrew Seal, Joseph Mfutso-Bengo, Martin Chalkley, Joseph Collins, Tara D Mangal, Pemphero N Mphamba, Rachel E Murray-Watson, John Phuka, Bingling She, Asif U Tamuri, Andrew Phillips, Paul Revill, Timothy B Hallett, Tim Colbourn

## Abstract

**Background:** Acute malnutrition remains a major public health challenge among children under five in Malawi due to undetected and untreated cases. While several policies and programmes are in place, they face significant resource input and implementation constraints. In this study, we evaluate the potential health impact and cost-effectiveness of three interventions designed to address constraints along the care pathway in childhood acute malnutrition management. These include improving early recognition of symptoms by caregivers, increasing attendance at routine growth monitoring visits through community outreach, and scaling up the availability of therapeutic food supplements.

**Methods and Findings:** We use a newly developed model representing the natural history and management of acute malnutrition, implemented within the *Thanzi La Onse* (TLO) dynamic individual-based simulation framework, which captures the public health system in Malawi. Each of the three interventions is assessed both individually and in combination, translated into seven scenarios which we evaluate in comparison to the status quo. The optimal strategy combines two interventions, improved caregiver awareness of early symptoms with increased availability of therapeutic food supplements. Over five years, this strategy is predicted to avert 840,470 (95% CI: 682,057–998,883) DALYs with total incremental costs of $34 million. This corresponds to an annual health expenditure increase of $0.32 per capita. At a cost-effectiveness threshold of $76 per DALY averted, the strategy results in an incremental net health benefit of 394,252 (95% CI: 235,839–552,665) DALYs averted.

**Conclusions:** The cost-effective strategy for addressing constraints in childhood acute malnutrition management is simultaneously improving caregiver recognition of early symptoms and expanding therapeutic food supplement availability. Out of the seven scenarios evaluated, this integrated approach was found to be the optimal strategy within the Malawian public health system, yielding substantial health at modest costs. These findings provide critical evidence to inform national policy and guide investment prioritisation for the management of childhood acute malnutrition.

## 1 Introduction

Malawi, a low-income country in Southern Africa, has made notable progress in reducing child mortality. Between 2016 and 2022, under-five mortality declined from 53.2 to 40.1 deaths per 1,000 live births [1]. Nevertheless, mortality rate remains significantly above the Sustainable Development Goal (SDG) 3.2 target [2]. This progress was achieved despite three-quarters of the population living below the international poverty line ($3.00 per day), extreme poverty affecting one in four, and livelihoods frequently disrupted by climate shocks and subsequent food insecurity [3,4]. These socioeconomic vulnerabilities contribute to persistent undernutrition among children. On the other hand, mortality reduction has not been matched by similar improvements in nutritional status, the prevalence of underweight, stunting, wasting, and overweight has remained stagnant [5,6]. This suggests that while Malawi has successfully addressed some primary causes of child mortality, socioeconomic vulnerabilities continue to exert a disproportionate influence on malnutrition, representing a distinct and persistent obstacle in child health.

Malnutrition is defined as the insufficient, excessive, or unbalanced intake of energy and nutrients needed to maintain good health [7]. The term malnutrition refers to all forms of deficiencies, excesses, or imbalances in a person’s intake of energy and/or nutrients, including undernutrition, micronutrient-related malnutrition, and overnutrition [8]. Undernutrition in children manifests in both chronic and acute forms. Chronic malnutrition, often indicated by stunting, reflects long-term nutritional deprivation. Acute malnutrition, the focus of this study, is a more immediate and life-threatening condition. It can be identified through indicators such as wasting—defined as weight-for-height (or weight-for-length in infants) less than two standard deviations below WHO Child Growth Standards [9]—, the presence of nutritional oedema, or, in children over six months of age, a mid-upper arm circumference (MUAC) below 125 mm. Based on severity, acute malnutrition is classified into moderate acute malnutrition (MAM) and severe acute malnutrition (SAM). National survey data show that the prevalence of moderate or severe wasting among children under five has remained largely unchanged, at 2.7% in 2015/16 (0.6% severe) and 2.6% in 2019/20 (0.7% severe) [5,6]. Although Malawi has already met the SDG 2.2 target of reducing wasting prevalence below 5% [10], the more pressing concern is the elevated risk of disability-adjusted life years (DALYs), primarily driven by mortality, associated with undetected or untreated acute malnutrition. Children with SAM are up to nine times more likely to die than well-nourished peers [7,11]. Survivors may face lifelong sequelae, including increased risks of cardiometabolic non-communicable diseases (NCDs), neurodevelopmental impairments, reduced physical strength, and mental health challenges [12–14].

Malawi has adopted several policies to reduce the burden of acute malnutrition. Growth monitoring visits are intended to detect early signs of malnutrition, incorporated with routine immunisation visits for infants, and via routine growth monitoring visits from one year of age. Children who seek care for any reason (related or unrelated) are indicated to also be assessed for acute malnutrition as part of the Integrated Management of Childhood Illness (IMCI) strategy [15]. Since 2012, the Community-Based Management of Acute Malnutrition (CMAM) programme has been implemented as part of Malawi’s Essential Health Package, with updated guidelines introduced in 2016 [16]. CMAM includes a supplementary feeding programme (SFP) for children with MAM, outpatient therapeutic care (OTC) for children with SAM without medical complications, and inpatient therapeutic care (ITC) for children with SAM involving medical complications.

However, these policies face significant implementation challenges. Detection often occurs late due to low attendance and poor recognition at growth monitoring and under-five clinic sessions [17], particularly after infancy when growth monitoring is no longer integrated with routine immunisation [18]. This is compounded by delayed care-seeking, as early signs of malnutrition are frequently unrecognised or perceived as insufficiently serious by caregivers and health care workers [19,20]. Moreover, the availability of therapeutic foods, such as Ready-to-Use Therapeutic Foods (RUTF) and Super Cereal Plus (CSB++), is heavily donor-dependent. This reliance on external funding has historically resulted in inconsistent domestic financing for nutrition supplies [21], contributing to frequent stockouts. Management is further exacerbated by sub-optimal diagnosis quality and a lack of long-term monitoring to confirm sustained recovery. These gaps contribute to higher relapse rates and treatment interruptions, increasing the risk that children with MAM deteriorate into SAM and, eventually, death.

Previous studies have largely concentrated on interventions addressing individual components of acute malnutrition management, such as prevention, SFP, OTC, ITC, or post-discharge, with limited attention to the implementation challenges that affect their outcomes [22–27]. This highlights the need for systems-based holistic analyses to better understand the potential of addressing these challenges and to inform strategic investment decisions in acute malnutrition management.

This study addresses that gap by applying a newly developed model representing the natural history and management of acute malnutrition, implemented within the *Thanzi La Onse* (TLO) dynamic simulation framework of the whole public health system in Malawi, which incorporates interactions among health conditions, demographic trends, care-seeking patterns, and resource constraints [28]. We evaluate the cost-effectiveness and health impacts of three interventions aimed at addressing the constraints in the management of acute malnutrition: (1) increasing attendance at routine growth monitoring visits through community outreach; (2) improving early recognition of acute malnutrition symptoms by caregivers; and (3) scaling up the availability of therapeutic food supplements. These interventions target the key bottlenecks in detection, care-seeking, and treatment supply, and are assessed both individually and in combination for their potential to avert DALYs in children under five.

## 2 Methods

This section details the modelling approach, outlining the structure of the *Thanzi La Onse* (TLO) health system simulation framework and the integration of the *Wasting* model, which simulates the natural history and clinical management of acute malnutrition. The natural history refers to the modelled progression and recovery of acute malnutrition in the absence of treatment. The management component includes detection and treatment pathways and outcomes. We also describe the constraints in management, the addressing of which is evaluated in this study. We then describe the modelled interactions between acute malnutrition and other health conditions within the TLO framework, followed by the calibration process. Finally, we present the intervention scenarios simulated and introduce the analysed outcomes.

More details can be found in S1 Appendix, including details regarding parameter values and their use in the model; consumables dispensed with treatments (amount, cost); parameters updated for scenarios; DALY weights attributed to health conditions related to acute malnutrition; and implementation costs calculations with sensitivity analysis.

### 2.1 Natural history and management of acute malnutrition within the TLO framework

This study employs the *Thanzi La Onse* (TLO) dynamic simulation framework of the Malawian health system, which integrates demographic and lifestyle factors, simulates the epidemiology of endemic diseases, and models individual care-seeking behaviour [29]. The framework facilitates the calculation of disability-adjusted life years (DALYs) and health system costs. While TLO can simulate constraints across personnel, equipment, medicines, and bed capacity [28,29], this study focuses specifically on medical consumable constraints. We assume other health system factors are not limiting, allowing us to isolate the impacts and costs that scale directly with treatment uptake, the improvement of which is the primary aim of the interventions evaluated in this study. This approach reflects clinical realities where severe cases are often prioritised regardless of formal bed capacity; furthermore, it avoids the uncertainties associated with current data limitations regarding the sub-national distribution of health workers and equipment.

The *Wasting* model tracks the acute malnutrition status and clinical trajectory of children under five through a set of individual attributes. These properties enable the model to monitor transitions between acute malnutrition states and the delivery of therapeutic care; a comprehensive list is provided in Table A1 in *Appendix A*. Associated model parameters are listed in Table A2 in *Appendix A*, with their implementation logic further described in *Appendix B*. We refer to ‘wasting’ as one of the physiological processes that often occurs during acute malnutrition and is determined for diagnostic purposes from the weight-for-height Z-score (WHZ), and to ‘acute malnutrition’ as the status given by diagnostic criteria based on the indicators (wasting, MUAC, and/or presence of nutritional oedema) as shown in Table 1 [30,31]. For the purposes of SAM treatment, SAM cases are further classified according to the presence or absence of medical complications. The specific complications are not explicitly modelled, only the presence or absence of “complications” is simulated.

**Table 1.**
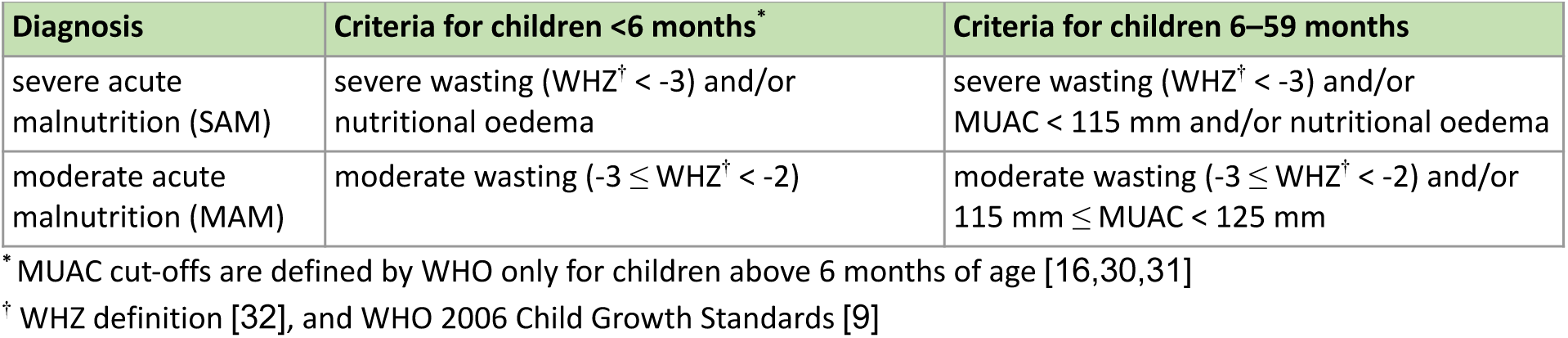
Acute malnutrition diagnostics criteria.

| Diagnosis | Criteria for children <6 months* | Criteria for children 6–59 months |
| --- | --- | --- |
| severe acute malnutrition (SAM) | severe wasting ($WHZ^{\dagger} < -3$) and/or nutritional oedema | severe wasting ($WHZ^{\dagger} < -3$) and/or $MUAC < 115$ mm and/or nutritional oedema |
| moderate acute malnutrition (MAM) | moderate wasting ($-3 \leq WHZ^{\dagger} < -2$) | moderate wasting ($-3 \leq WHZ^{\dagger} < -2$) and/or $115 \text{ mm} \leq MUAC < 125$ mm |

\* MUAC cut-offs are defined by WHO only for children above 6 months of age [16,30,31]
<sup>†</sup> WHZ definition [32], and WHO 2006 Child Growth Standards [9]

The ‘Natural History’ of acute malnutrition describes the modelled disease evolution in the absence of treatment (Fig 1). Progression is defined by the severity of wasting, categorised by the weight-for-height z-score (WHZ) to account for limited data on MAM and SAM prevalence and incidence. Natural recovery is illustrated through transitions between acute malnutrition states (SAM, MAM, or well-nourished) to align with treatment recovery pathways, whereby individuals transition into the adjacent, less severe health category. Mortality is modelled exclusively for children meeting SAM diagnostic criteria (Table 1). Under this framework, children in the moderate or severe wasting categories face mortality risk if they meet any qualifying SAM criteria (WHZ, MUAC, or oedema). In the absence of treatment, younger children are assigned a higher relative risk of death [33]. The incidence of new wasting cases is determined monthly and depends on several risk factors, including age, size at birth, preterm birth, and household wealth. The duration of untreated wasting episodes is simulated as 81 days for moderate, and 45 days for severe wasting [34]. Further details on the modelling ‘Natural History’ are provided in Fig B1 in *Appendix B*.

**Fig 1.**
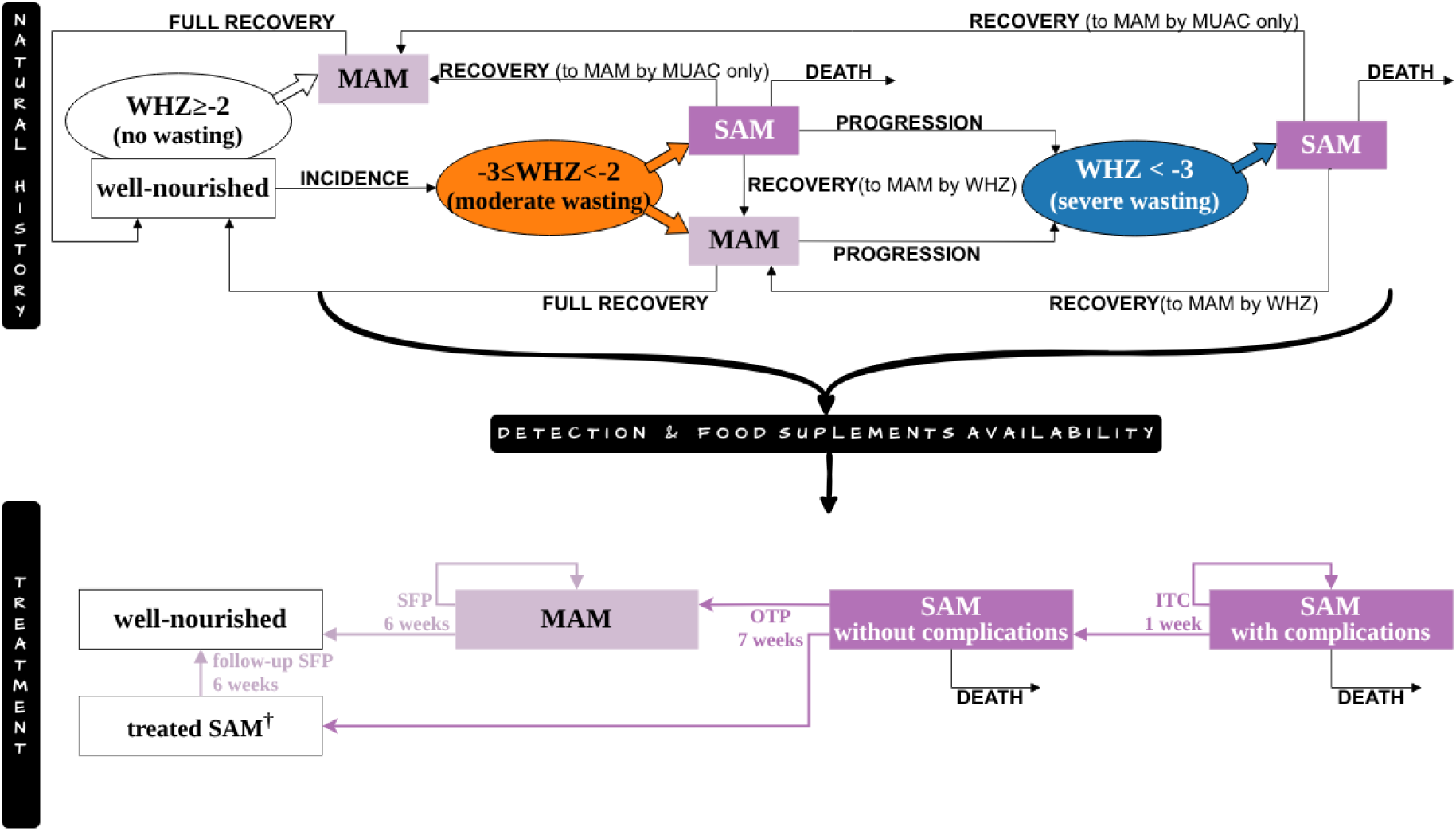
Conceptual overview of the modelled natural history and treatment of acute malnutrition. ^†^’treated SAM’ refers to children who have fully recovered from SAM and require follow-up care through a SFP

Engagement with the health system may shift a child’s trajectory from the ‘Natural History’ to the ‘Treatment’ pathway of the model (Fig 1). Detection occurs either through symptomatic care-seeking at any facility level or during routine growth monitoring at primary care facilities. Care-seeking is driven by the TLO framework, which accounts for sex, age, wealth, and residence [35–37]. Under the status quo, we assume full caregiver awareness of SAM symptoms, resulting in a high probability of care-seeking (over 90%). In contrast, care-seeking is not assumed for MAM specifically; however, because malnutrition screening is integrated into the Integrated Management of Childhood Illness (IMCI) guidelines [15], any child seeking care for any reason (whether related to acute malnutrition or not) may be detected. We represent this by modelling the proportion of cases expected to be identified if screened, based on reported data which implicitly account for real-world diagnostic failures and misdiagnoses. Additionally, as part of the Malawi Community-based Management of Acute Malnutrition (CMAM) programme [16], routine growth monitoring serves as a detection point. Attendance frequency and probability vary by age group, with scheduled visits becoming less frequent as age increases [35]; attendance is highest among children aged 0–11 months and lowest among those aged 12–23 months [38,39]. If acute malnutrition is diagnosed, the child is referred for appropriate treatment. We model admission as being contingent on the availability of food supplements; this reflects the reality that clinical benefits are only realised when therapeutic supplies are present. Consequently, cases where supplements are unavailable are modelled as remaining on the ‘Natural History’ trajectory, as they would not receive the intended treatment effect. Conversely, where supplies are available, admission to treatment supersedes the ‘Natural History’ trajectory with outcomes defined by the ‘Treatment’ pathway (Fig 1).

Following Malawi guidelines for CMAM [16], the model incorporates three treatment modalities. The duration of each was estimated based on existing literature [16,40–42]:

- SFP (MAM): CSB++ administered over six weeks.
- OTP (SAM without complications): Recommended medicines and RUTF administered over seven weeks.
- ITC (SAM with complications): Hospital-based care, including recommended medicines, stabilisation (F-75 milk), and a transition phase (RUTF) over one week.

Treatment outcomes (recovery, re-enrolment, or mortality) and follow-up care are illustrated in the ‘Treatment’ section of Fig 1. Re-enrolment and follow-up both depend on supplement availability, which reflects seasonal variations at primary (SFP/OTP) and secondary (ITC) facility levels [43]. The average monthly probabilities of treatment availability are presented in Fig 2. Detailed modelling parameters and logic are provided in *Appendix B* (subsection *Treatment*) and illustrated in Fig B2.

**Fig 2.**
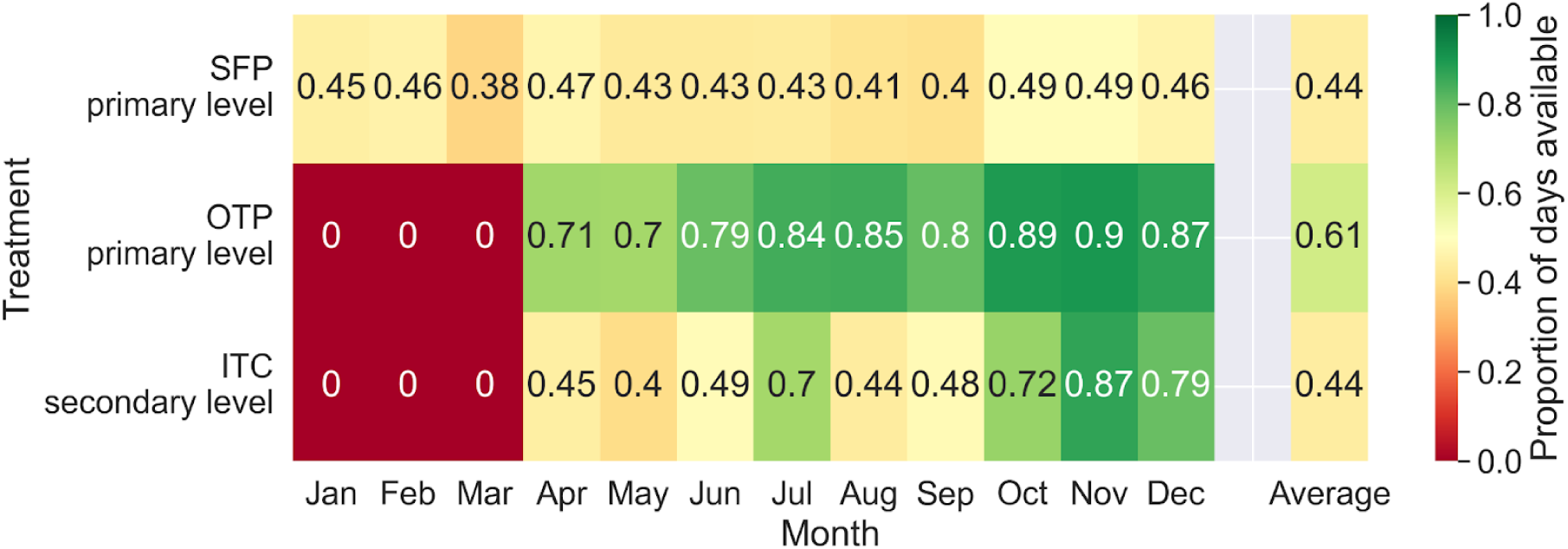
Monthly average availability of treatments at corresponding facility level. Average availability of treatments is based on average availability of the food supplements used for the treatment, CSB++ for SFP, RUTF for OTP, and both F-75 therapeutic milk and RUTF for ITC (Fig B3 in *Appendix B*), that were estimated and calibrated, where feasible, using data availability, as detailed in [44]. The average availability of treatments is calculated as the product of the average availability probabilities of food supplements used for the treatment.

Acute malnutrition interacts with multiple health conditions. SAM is a major risk factor for mortality from infectious diseases, including acute lower respiratory infection (ALRI) and diarrhoea. Children aged 2–59 months with SAM have seven times higher odds of dying due to untreated ALRI compared to those with MAM or well-nourished [45], and intravenous antibiotic treatment for severe pneumonia is twice as likely to fail in SAM and 1.5 times more likely in MAM cases [46] (modelled in the *ALRI* model of TLO framework [47]). Diarrhoea-related mortality is four times higher in children with SAM [48], with increased risks of incidence (1.2 times) [49] and progression to persistent diarrhoea (1.7 times) [50] (*Diarrhoea* model [51]). A history of wasting raises the risk of stunting incidence or progression by 1.9 times [52] (*Stunting* model [53]). Stunting is assumed not to cause direct deaths or incur years lived with disability (YLDs), which is consistent with the Global Burden of Disease Study 2019 (GBD 2019) [54]. However, the impact of stunting manifests indirectly, severely stunted children have a 33.3 times higher risk of prolonged diarrhoea becoming persistent [55] (*Diarrhoea* model). Conversely, children born small for gestational age (SGA) and/or preterm have elevated risks of wasting, 3.1 times higher for SGA and preterm (24–36 weeks), 2.1 times for SGA at term, and 2.1 times for average gestational age but preterm [56] (*Wasting* model).

### 2.2 Calibration

A five-year burn-in period (2010–2014) was applied to allow the model to stabilise. Model calibration was conducted in two stages. First, age-specific relative risks of monthly moderate wasting incidence and monthly progression to severe wasting were calibrated using simplified ordinary differential equations (ODE), see *Appendix C* for more details. Second, simulation-based calibration was conducted to estimate magnitude of these risks, the baseline risks of monthly moderate wasting incidence and progression to severe wasting for the reference group of children (i.e. under five months of age), as well as baseline risks of death due to untreated SAM—assuming a 0.8-fold lower risk of death in children aged one year and older—and death due to SAM when treated with outpatient or inpatient care, assuming equalised risk across ages under treatment. See *Appendix B* for the calibrated risks, and *Appendix A* for their translation into parameter values. The age-specific proportions of moderate and severe wasting in 2016 were calibrated to DHS 2015–2016 data [5], and in 2020 to MICS 2019–2020 data [6]. Annual direct deaths averaged over 2015–2019 were calibrated to deaths due to ‘protein-energy malnutrition’ as reported in GBD 2019 [54], see Fig 3.

**Fig 3.**
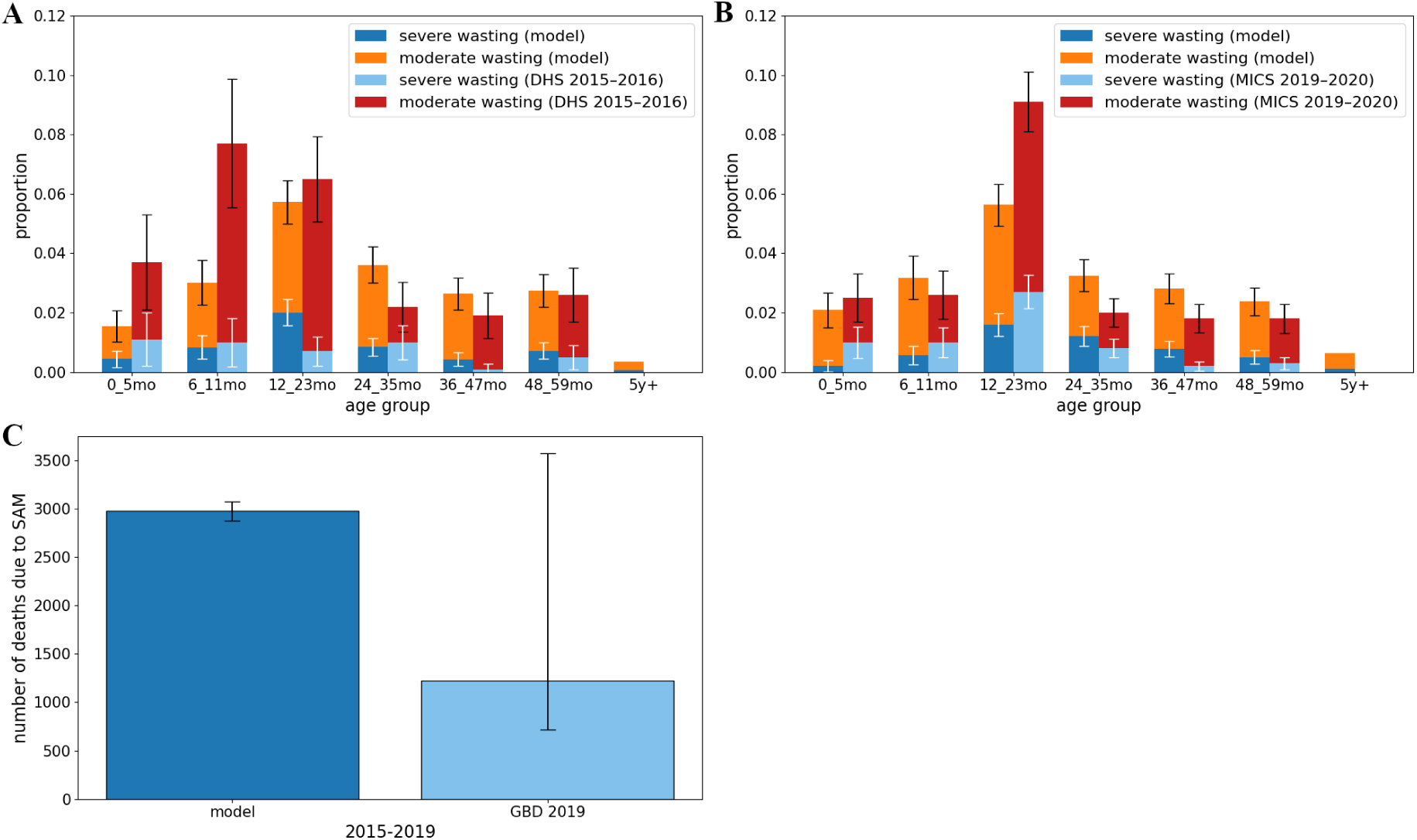
Calibrated outcomes for children under five. Wasting prevalence by age group in **(A)** 2016; **(B)** 2020; uncertainty bounds represent 95% CIs for severe wasting (white) and any wasting (black). **(C)** Average annual direct deaths due to severe acute malnutrition over the period 2015–2019; means and 95% CIs.

### 2.3 Scenarios

We model combinations of the three interventions in eight scenarios to evaluate the potential impact of interventions addressing constraints in childhood acute malnutrition management in Malawi. The Status Quo (SQ) reflects current conditions, including existing growth monitoring attendance, no caregiver awareness of MAM symptoms, and default food supplement availability, corresponding to parameter values as listed in Table A2 in *Appendix A*. The other seven scenarios simulate the implementation of an individual intervention or their combination starting in 2026: the ‘Growth Monitoring’ intervention assumes full attendance at growth monitoring visits for children aged 1–5 years (increased attendance at routine visits once monitoring is no longer integrated with immunisation); the ‘Care-Seeking’ intervention assumes 100% caregiver awareness of MAM symptoms; and the ‘Food Supplements’ intervention assumes full availability of all therapeutic food supplements (CSB++, RUTF, F-75 milk) across all months of the year. The overview of all eight scenarios including corresponding changes to parameters are shown in Table B4 in *Appendix B*. While the Growth Monitoring or Care-Seeking interventions are not directly focusing on increase of Food Supplements, the default assumption of probability of treatment availability (Fig 2) is preserved when Food Supplements intervention is not implemented along with them, meaning the availability of food supplements had to be adjusted proportionally to the change in demand for the treatment resulting from these interventions. Each scenario is simulated for the period from 1 January 2010 to 31 December 2025 under the status quo assumptions, with corresponding interventions modelled as implemented from 1 January 2026 to 31 December 2030. The TLO framework is run using a simulated cohort of 100,000 individuals representative of the Malawian population, with outputs scaled to the national population. To account for uncertainty due to stochasticity, each scenario is executed using 30 distinct random seeds.

### 2.4 Health outcomes, costs, and cost-effectiveness

We analysed the number of disability-adjusted life years (DALYs) averted during 2026–2030 compared with the SQ, calculated as the difference in cumulative DALYs between each intervention scenario and SQ for each run. Means and 95% confidence intervals (CIs) across all runs are presented as results. DALYs were computed as the sum of years of life lost and years lived with disability (attributed to any of the health conditions incorporated in the TLO framework) [57], using monthly disability weights based on GBD 2019 [54]. Because acute malnutrition increases the risk of ALRI and diarrhoea, the cause-specific averted DALYs attributable to SAM, ALRI, and diarrhoea are reported in *Appendix D*.

Modelled costs included medical consumables, wastage, and supply chain expenses, presented in 2023 USD [44]. The treatment is administered only if all essential consumables, the food supplements, are available. The cost of treatment is determined by the specific supplements and other consumables administered (depending on their availability). The cost per case is $43 for SFP, $45–$47 for OTP, and $5–$7 for ITC. When accounting for the full treatment pathway, specifically the important transition from ITC to OTP and subsequent MAM follow-up, the total cumulative costs per case are $43 for MAM, $88–$89 for uncomplicated SAM, and $93–$97 for complicated SAM, see *Appendix B* (subsection *Consumables*) for details. The additional implementation costs were estimated from a Ministry of Health (MoH) perspective using a unit cost of $0.53 in the first year (to cover start-up costs) and $0.37 in subsequent years per targeted individual (the parent of a newborn for Growth Monitoring and Care-Seeking, or the malnourished child for Food Supplements) annually (2023 USD) for Malawi [58–61]. Partial operational cost-sharing (50% of the additional implementation costs [62]) was assumed for joint implementation of Growth Monitoring and Care-Seeking, reflecting delivery synergies. Details on how these values were determined are available in *Appendix B* (subsection *Costs and Sensitivity analysis*).

Incremental cost-effectiveness analysis was conducted to compare each alternative, identifying non-dominated strategies to define the cost-effectiveness frontier. This analysis is based on the Malawi-specific cost-effectiveness threshold (CET) of $76 per DALY averted. This threshold was derived from the $61 (2016 USD) identified as the threshold for affordability for the Malawi Essential Health Package [63], inflated to 2023 USD [64]. We report total and incremental costs, DALYs averted, and the Incremental Cost-Effectiveness Ratio (ICER) per DALY averted relative to the next less costly non-dominated strategy. To estimate the aggregate effect on population health, we calculated the Incremental Net Health Benefit (INHB) compared to the SQ [65]. Furthermore, to identify the maximum allowable total costs while a scenario remains cost-effective, we determined the Incremental Net Monetary Benefit (INMB) as:

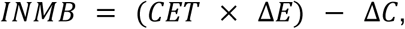

where Δ*E* represents the incremental DALYs averted, and Δ*C* the total incremental costs compared to the Status Quo, and *CET* is the cost-effectiveness threshold. The maximum allowable total costs are identified as the point where the monetised health benefits are entirely offset by the total costs (INMB = 0), with values provided in *Appendix B* (subsection *Costs and Sensitivity analysis*).

All costs are presented in 2023 USD. All costs and health outcomes were discounted at 3% annually [66].

### 2.5 Sensitivity analysis

Three-way sensitivity analysis considered varied unit cost, proportion of shared implementation costs when Growth Monitoring and Care-Seeking interventions are implemented jointly, and a multiplier of unit cost assumed for Food Supplements intervention, as detailed in *Appendix B* (subsection *Costs and Sensitivity analysis*).

### 2.6 Ethics statement

The *Thanzi La Onse* project received ethical approval from the *College of Medicine Malawi Research Ethics Committee* (COMREC, P.10/19/2820) in Malawi. Only publicly available anonymised secondary data is used in the *Thanzi La Onse* modelling framework; therefore, individual informed consent was not required.

## 3 Results

When implementing a single intervention, the Food Supplements intervention results in the greatest estimated impact, averting 604,966 (95% CI: 421,915–788,017) DALYs during 2026–2030 compared to the Status Quo. In contrast, the Care-Seeking and Growth Monitoring interventions implemented alone show no statistically significant effects, with mean averted DALYs estimated at approximately 20% and 13% of the impact achieved by the Food Supplements intervention, respectively (Table 2).

**Table 2.** Cost-effectiveness metrics.

| Scenario | Total incremental costs<sup>†</sup> vs SQ (2023 USD) | DALYs averted vs SQ (95% CI) | Incremental costs vs less costly strategy (2023 USD) | Incremental DALYs averted vs less costly strategy | ICER per DALY averted vs less costly strategy (2023 USD) | Incremental net health benefit vs SQ (95% CI) |
| --- | --- | --- | --- | --- | --- | --- |
| *Status Quo* | 0 | 0 | n/a | n/a | n/a | 0 |
| *Care-Seeking* | 3,580,169 | 123,859<br>(-55,953; 303,670) | 3,580,169 | 123,859 | 29 | 76,751<br>(-103,060; 256,562) |
| *Growth Monitoring* | 4,842,883 | 75,984<br>(-33,745; 185,713) | n/a | n/a | dominated | 12,262<br>(-97,467; 121,991) |
| *Growth Monitoring and Care-Seeking* | 6,992,232 | 228,055<br>(74,384; 381,727) | 3,412,063 | 104,196 | 33 | 136,052<br>(-17,619; 289,724) |
| *Care-Seeking and Food Supplements* | 33,912,588 | 840,470<br>(682,057; 998,883) | 26,920,356 | 612,415 | 44 | 394,252<br>(235,839; 552,665) |
| *Growth Monitoring and Care-Seeking and Food Supplements* | 36,056,207 | 846,482<br>(722,700; 970,264) | 2,143,619 | 6,012 | 357 | 372,058<br>(248,276; 495,841) |
| *Growth Monitoring and Food Supplements* | 37,447,616 | 785,748<br>(643,800; 927,696) | n/a | n/a | dominated | 293,016<br>(151,068; 434,964) |
| *Food Supplements* | 40,935,570 | 604,966<br>(421,915; 788,017) | n/a | n/a | dominated | 66,340<br>(-116,711; 249,391) |

Notes: Scenarios highlighted with a light green background are non-dominated and lie on the cost-effectiveness frontier (Fig 4). Values represent means across 30 stochastic runs; the 95% CIs reflect model stochasticity.
<sup>†</sup> Total incremental costs represent the sum of incremental consumable-related and additional implementation costs. For a detailed breakdown of cost categories and the calculated 'Maximum allowable total costs' (i.e., budgetary headroom), see Fig B4 and Table B6 in *Appendix B* (subsection *Costs and Sensitivity analysis*).

Implementing multiple interventions simultaneously achieves a greater impact, as combined approaches always yield better results than any subset implemented in isolation. Notably, the health impact is substantially strengthened when any intervention or combination is supported by the Food Supplements intervention. Specifically, as shown in Table 2, when complemented with Food Supplements, the impact of Growth Monitoring multiplies tenfold (from 75,984 to 785,748 DALYs averted), of Care-Seeking sevenfold (from 123,859 to 840,470 DALYs averted), and of the combined Growth Monitoring and Care-Seeking scenario approximately fourfold (from 228,055 to 846,482 DALYs averted). All scenarios that include Food Supplements, particularly the combination of all three interventions, achieve high overall effectiveness. The maximum effect, reached when all three interventions are implemented, results in averting 846,482 (95% CI: 722,700–970,264) DALYs (Table 2).

The cost-effectiveness analysis identified four strategies on the cost-effectiveness frontier (Fig 4). The incremental investment pathway reveals the optimal choice at CET of $76. Moving from the Status Quo, the initial step is the Care-Seeking scenario with an ICER of $29 per DALY averted. Stepping up, the scenario combining Growth Monitoring and Care-Seeking results in an ICER of $33 per additional DALY averted. Combining Care-Seeking with Food Supplements remains cost-effective, at an incremental cost of $46 per additional DALY averted and provides the largest beneficial effect on population health as represented by INHB of 394 thousand DALYs averted. However, further expanding this intervention package to include Growth Monitoring is not cost-effective, as the incremental investment required ($357 per additional DALY averted) significantly exceeds the CET (Table 2, and Fig 4).

**Fig 4.**
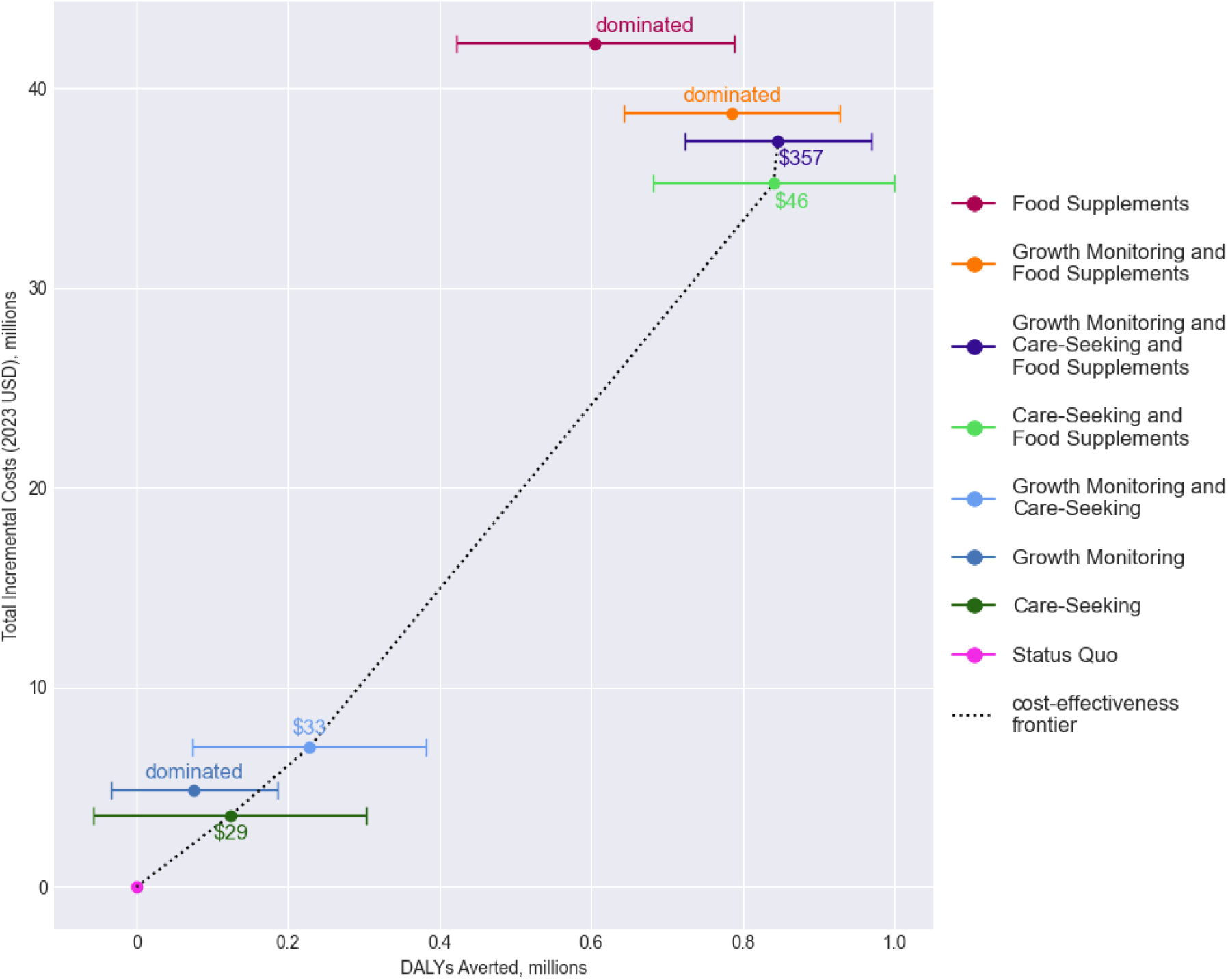
Cost-effectiveness plane.

Scenarios are ordered by increasing total incremental costs, and those identified as non-dominated strategies on the cost-effectiveness frontier (Fig 4) are highlighted (light green background) in Table 2. At the $76/DALY cost-effectiveness threshold, the Care-Seeking and Food Supplements scenario is identified as the optimal strategy, yielding the greatest incremental net health benefit and estimated to avert 840,470 DALYs (95% CI: 682,057–998,883) (Table 2). The total incremental costs for this strategy are estimated at $34 million over the five-year intervention period, equivalent to an annual per capita investment of approximately $0.32 [67], or 0.8 per cent of the most recent per capita health expenditure estimate of $41.09 (inflated to 2023 USD) [68,64]. The INMB, calculated at $29,963,157 (95% CI: 17,923,759–42,002,554) (not shown), indicates substantial budgetary headroom, as the scenario remains cost-effective up to total incremental costs of $63,875,745 (95% CI: 51,836,347–75,915,143) (Table B6 in *Appendix B*, subsection *Costs and Sensitivity analysis*). This allowable cost threshold corresponds to an annual per capita expenditure requirement of $0.61, or 1.5 per cent of current per capita health spending.

Three-way sensitivity analysis (Fig B5 in *Appendix B*, subsection Costs and Sensitivity analysis) indicates that while findings are qualitatively robust to variations in shared implementation costs for Growth Monitoring and Care-Seeking and to the assumed proportion of unit cost for the Food Supplements intervention, the results are sensitive to the absolute unit cost. Specifically, when applying a substantially higher baseline unit cost ($9.17 [69]), cost-effectiveness is compromised. Under this high-cost scenario, the Status Quo becomes the optimal strategy if the unit cost for Food Supplements exceeds $12.8 per malnourished child (corresponding to 1.4 times the baseline unit cost, the FS multiplier in sensitivity analysis); otherwise, the Food Supplements scenario emerges as the sole cost-effective option at the CET of $76.

Additionally, cause-specific analysis revealed that the majority of averted DALYs are attributable to reductions in SAM, with no observable overall effect on ALRI or diarrhoea morbidity and mortality (*Appendix D*, section *Cause-specific outcomes*).

While the interventions do reduce deaths due to these conditions, the limited overall indirect impact is explained by the low baseline prevalence of SAM among children dying from ALRI (3.8%) and diarrhoea (5.4%) in Malawi (*Appendix D*, section *Cause-specific outcomes*). This is due to the higher under-five prevalence of ALRI and diarrhoea [70,71] relative to wasting [6], suggesting that while SAM increases individual vulnerability, it is not the predominant underlying driver of most respiratory or diarrhoeal mortality.

## 4 Discussion

Our cost-effectiveness analysis, which focused on addressing three implementation constraints within Malawi’s acute malnutrition management system, identified the combination of improved caregiver awareness of symptoms in moderate cases of acute malnutrition leading to care-seeking (Care-Seeking intervention) and ensuring a reliable supply of food supplements (Food Supplements intervention) as the optimal scenario. This integrated approach (Care-Seeking and Food Supplements scenario) substantially improves health outcomes at modest costs, equivalent to an annual investment of only 0.8 per cent of per capita health expenditure. After accounting for the resources required to fund the programme, this scenario yields a net gain of 394,252 DALYs (INHB; Table 2), representing the greatest overall improvement in population health among all evaluated strategies. Furthermore, the economic value of these health gains is nearly double the implementation cost, providing a net value to Malawi’s population health of approximately $30 million over five years (INMB).

While acute malnutrition is a recognised risk factor for infectious diseases [45,46,48,49], we did not observe significant overall reductions in ALRI or diarrhoea morbidity. This limited indirect impact is likely due to the low baseline overlap between SAM and these conditions in Malawi, where ALRI and diarrhoea prevalence [70,71] significantly exceeds wasting prevalence [6]. These findings suggest that malnutrition management is not a substitute for targeted infectious disease interventions, both must be pursued in tandem. Future research should further explore these interactions, including impacts on vaccine effectiveness and other comorbidities such as malaria, measles, or tuberculosis [72–76].

A key strength of this work is the use of the comprehensive *Thanzi La Onse* (TLO) individual-based simulation framework [28]. Calibrated to local data, the framework realistically represents public health system behaviour and resource constraints, enabling a robust cost-effectiveness analysis that integrates both costs and health impacts across multiple interconnected conditions. However, our study explored the theoretical potential of fully addressing constraints in acute malnutrition management (increasing detection by assuming early symptoms are recognised by caregivers in all cases, achieving full attendance at routine growth monitoring, and ensuring a sufficient supply of food supplements to treat all patients). Future research should focus on analysing realistic, incremental improvements to enhance policy relevance.

To improve accuracy, future model iterations should address several critical data and structural limitations. Firstly, the current model does not explicitly model diagnostic failures, such as misdiagnoses due to missing equipment (e.g. MUAC tape or stadiometer) [77,78], measurement errors [79], or omitted measurements [34]. While these failures are implicitly captured within the observed prevalence and incidence data used for calibration, explicit modelling is required to evaluate interventions that aim to improve diagnostic accuracy or that may inadvertently alter misdiagnosis rates. Secondly, a further gap lies in the precise health workforce requirements for expanded treatment delivery. While the TLO framework is capable of simulating healthcare worker time-costs, empirical data are needed to model system responses to demand fluctuations—whether through reallocation from other services, overwhelming existing staff capacity, or treatment delays. Finally, the current version of the framework does not incorporate the long-term sequelae of acute malnutrition, such as cardiometabolic NCDs, neurocognitive deficits, and mental health problems that manifest in later life [12–14]. Whilst the focus on DALYs may not fully capture the impairment in educational and occupational attainment possibly resulting from neurocognitive deficits [80], this perpetuating cycle of vulnerability could be represented in future iterations by modelling intergenerational feedback loops. In such a framework, improved cognitive outcomes would influence household wealth, one of the determinants of wasting incidence, thereby affecting the risk for future generations.

The implications of these omissions are two-fold. Omitting explicit diagnostic and workforce constraints may lead to over-optimistic implementation of the scenarios. Diagnostic failures can diminish the effectiveness of detection-focused interventions or inflate costs through the over-treatment of less severe cases. Workforce constraints could increase recruitment costs or reduce broader system performance—including increased misdiagnosis rates—if staff are overwhelmed or diverted from other services. Conversely, the exclusion of long-term sequelae suggests our results are likely conservative, as their inclusion would significantly increase the DALYs averted by successful treatment. The TLO framework is well-suited to addressing these gaps in future iterations. Its architecture is already designed to simulate constraints across equipment and personnel [28,29] and can readily incorporate these mechanisms as sub-national data become available. Additionally, because the framework simulates health across the entire life-course, it is ideally placed to evaluate lifetime health impacts and potential feedback loops driven by improved social determinants once these long-term sequelae are parameterised and calibrated [81].

While incremental costs related to medical consumables are rigorously analysed [44], the uncertainty surrounding estimates of implementation costs (personnel, management, materials, diagnostic tools) must be acknowledged. Sensitivity analysis showed outcomes are sensitive to the unit cost assumed; however, comparison with the latest expenditure data on nutrition projects suggests our primary unit cost estimate is accurate. The higher unit cost of $9.17 serves as a conservative upper bound; despite efforts to exclude societal-perspective costs, this figure likely retains elements that would not be borne by the Ministry of Health. Ultimately, final policy decisions require precise budget and unit cost estimates before implementation.

Our findings provide a strategic evidence base for optimising acute malnutrition management in Malawi. To maximise population health, investment must prioritise an integrated strategy that concurrently addresses caregiver awareness and food supplements supply. This approach yields health gains that substantially outweigh the required financial investment. Furthermore, the underlying model serves as a durable decision-support tool, capable of rapidly re-evaluating cost-effectiveness as unit costs and national scaling priorities evolve.

## Supporting information

S1 Appendix. Supplementary appendix.

## Data availability statement

The *Thanzi La Onse* modelling framework is open source and available for review and usage at https://github.com/UCL/TLOmodel. In particular, the outputs analysed in this study can be reproduced from the tag accessible at https://github.com/UCL/TLOmodel/releases/tag/Janouskova_etal2026_wasting_inter <u>vs_v0.1</u>, using the scenario files from src/scripts/wasting_analyses/scenarios/100K. The scripts used to generate the plots in the manuscript and the appendix (heatmaps_cons_wast.py, calib_analysis_wasting.py, run_interventions_analysis_wasting.py) can be found in the src/scripts/wasting_analyses directory.

## Acknowledgement

This work was supported by the UK Research and Innovation as part of the Global Challenges Research Fund under Grant MR/P028004/1 and the Wellcome Trust under Grant 223120/Z/21/Z. TDM, REM, BS, TBH acknowledge funding from the MRC Centre for Global Infectious Disease Analysis (reference MR/R015600/1), jointly funded by the UK Medical Research Council (MRC) and the UK Foreign, Commonwealth & Development Office (FCDO), under the MRC/FCDO Concordat agreement and is also part of the EDCTP2 programme supported by the European Union.

## Author contributions

**Conceptualisation:** Eva Janoušková, Ines Li Lin, Tim Colbourn

**Data Curation:** Eva Janoušková, Ines Li Lin

**Formal Analysis:** Eva Janoušková

**Funding Acquisition:** Timothy B Hallett, Dominic Nkhoma, Paul Revill, Andrew Phillips, Tim Colbourn

**Investigation:** Eva Janoušková, Emilia Connolly, Dominic Nkhoma

**Methodology:** Eva Janoušková, Ines Li Lin, Sakshi Mohan, Andrew Seal, Andrew Phillips, Paul Revill, Timothy B Hallett, Tim Colbourn

**Project Administration:** Eva Janoušková, Tim Colbourn

**Resources:** Asif U Tamuri

**Software:** Eva Janoušková, Ines Li Lin, Emmanuel Mnjowe, Watipaso Mulwafu, Asif U Tamuri, Timothy B Hallett

**Supervision:** Tim Colbourn

**Validation:** Eva Janoušková

**Visualisation:** Eva Janoušková

**Writing – Original Draft Preparation:** Eva Janoušková

**Writing – Review & Editing:** Eva Janoušková, Ines Li Lin, Emmanuel Mnjowe, Watipaso Mulwafu, Emilia Connolly, Sakshi Mohan, Dominic Nkhoma, Andrew Seal, Joseph Mfutso-Bengo, Martin Chalkley, Joseph Collins, Tara D Mangal, Pemphero N Mphamba, Rachel E Murray-Watson, John Phuka, Bingling She, Asif U Tamuri, Andrew Phillips, Paul Revill, Timothy B Hallett, Tim Colbourn

## Supplementary Appendix

### Appendix A

#### Properties

Individual properties (<u>Table A1</u>) are stored during the simulation that carry the information on the history and current state of acute malnutrition.

**Table A1.**
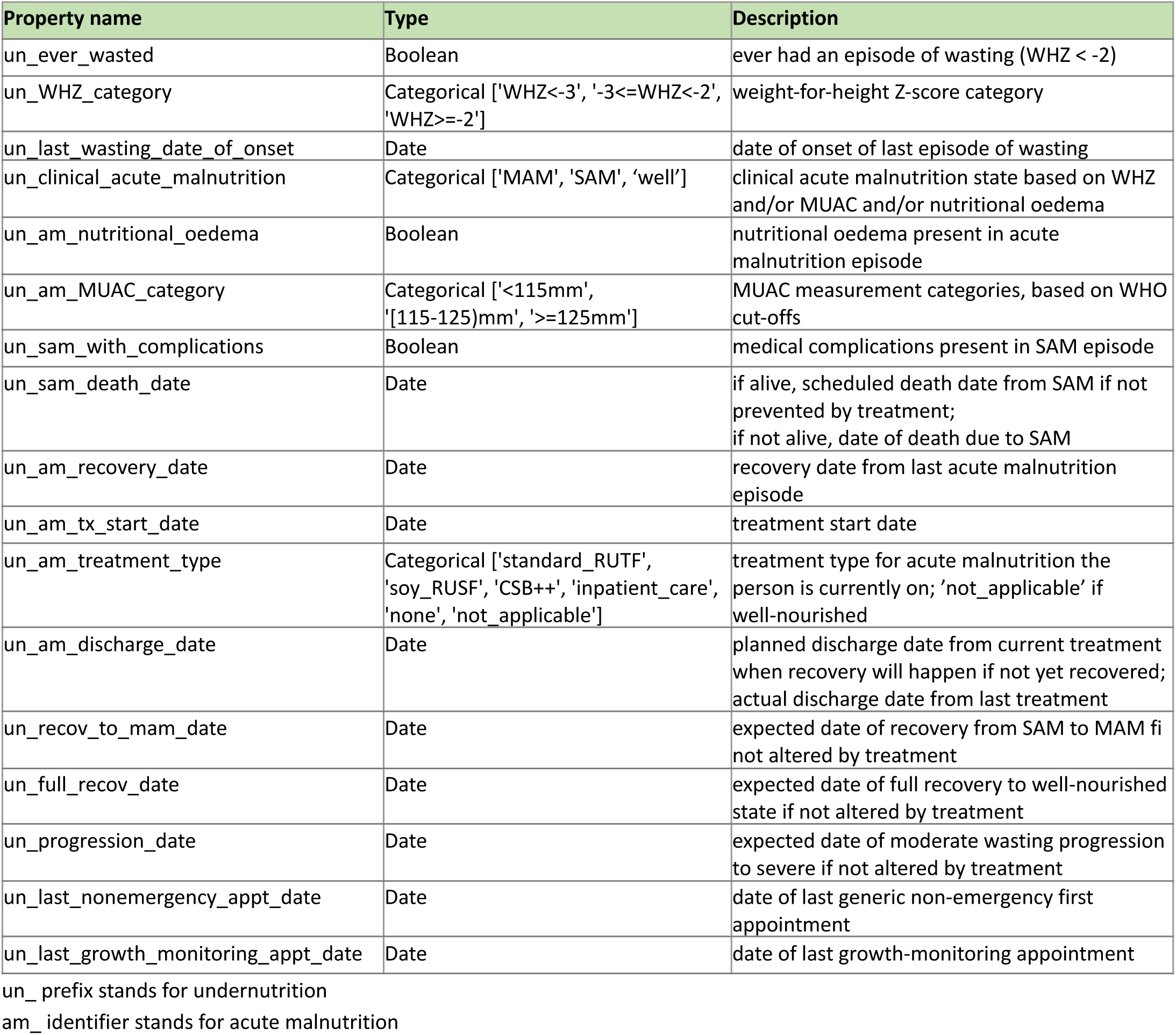
Properties of each individual related to the *Wasting* model.

| Property name | Type | Description |
| --- | --- | --- |
| un_ever_wasted | Boolean | ever had an episode of wasting (WHZ < -2) |
| un_WHZ_category | Categorical ['WHZ<-3', '-3<=WHZ<-2', 'WHZ>=-2'] | weight-for-height Z-score category |
| un_last_wasting_date_of_onset | Date | date of onset of last episode of wasting |
| un_clinical_acute_malnutrition | Categorical ['MAM', 'SAM', 'well'] | clinical acute malnutrition state based on WHZ and/or MUAC and/or nutritional oedema |
| un_am_nutritional_oedema | Boolean | nutritional oedema present in acute malnutrition episode |
| un_am_MUAC_category | Categorical ['<115mm', '[115-125)mm', '>=125mm'] | MUAC measurement categories, based on WHO cut-offs |
| un_sam_with_complications | Boolean | medical complications present in SAM episode |
| un_sam_death_date | Date | if alive, scheduled death date from SAM if not prevented by treatment;<br>if not alive, date of death due to SAM |
| un_am_recovery_date | Date | recovery date from last acute malnutrition episode |
| un_am_tx_start_date | Date | treatment start date |
| un_am_treatment_type | Categorical ['standard_RUTF', 'soy_RUSF', 'CSB++', 'inpatient_care', 'none', 'not_applicable'] | treatment type for acute malnutrition the person is currently on; 'not_applicable' if well-nourished |
| un_am_discharge_date | Date | planned discharge date from current treatment when recovery will happen if not yet recovered;<br>actual discharge date from last treatment |
| un_recov_to_mam_date | Date | expected date of recovery from SAM to MAM if not altered by treatment |
| un_full_recov_date | Date | expected date of full recovery to well-nourished state if not altered by treatment |
| un_progression_date | Date | expected date of moderate wasting progression to severe if not altered by treatment |
| un_last_nonemergency_appt_date | Date | date of last generic non-emergency first appointment |
| un_last_growth_monitoring_appt_date | Date | date of last growth-monitoring appointment |

un\_ prefix stands for undernutrition
am\_ identifier stands for acute malnutrition

#### Parameters

**Table A2.**
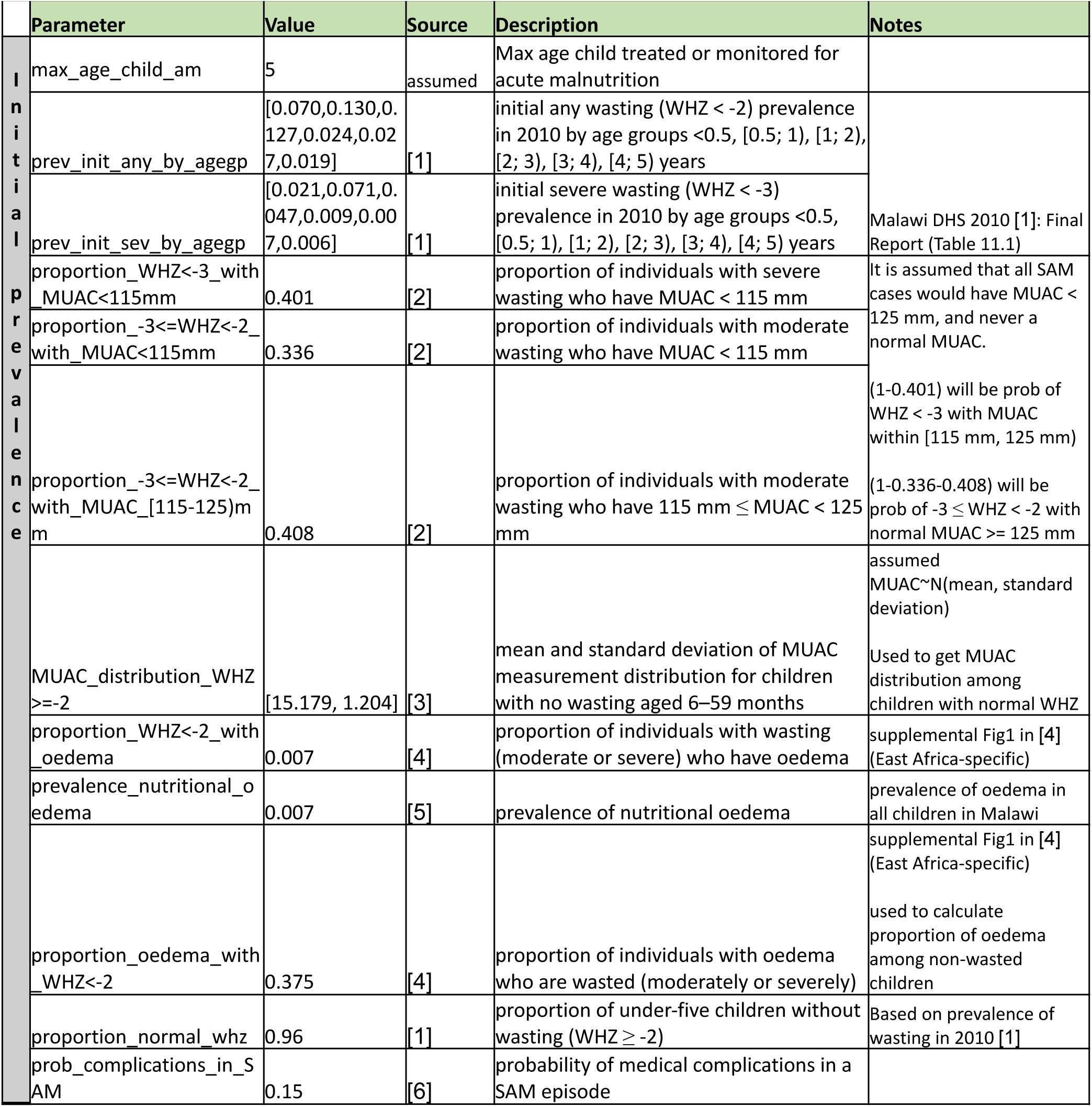
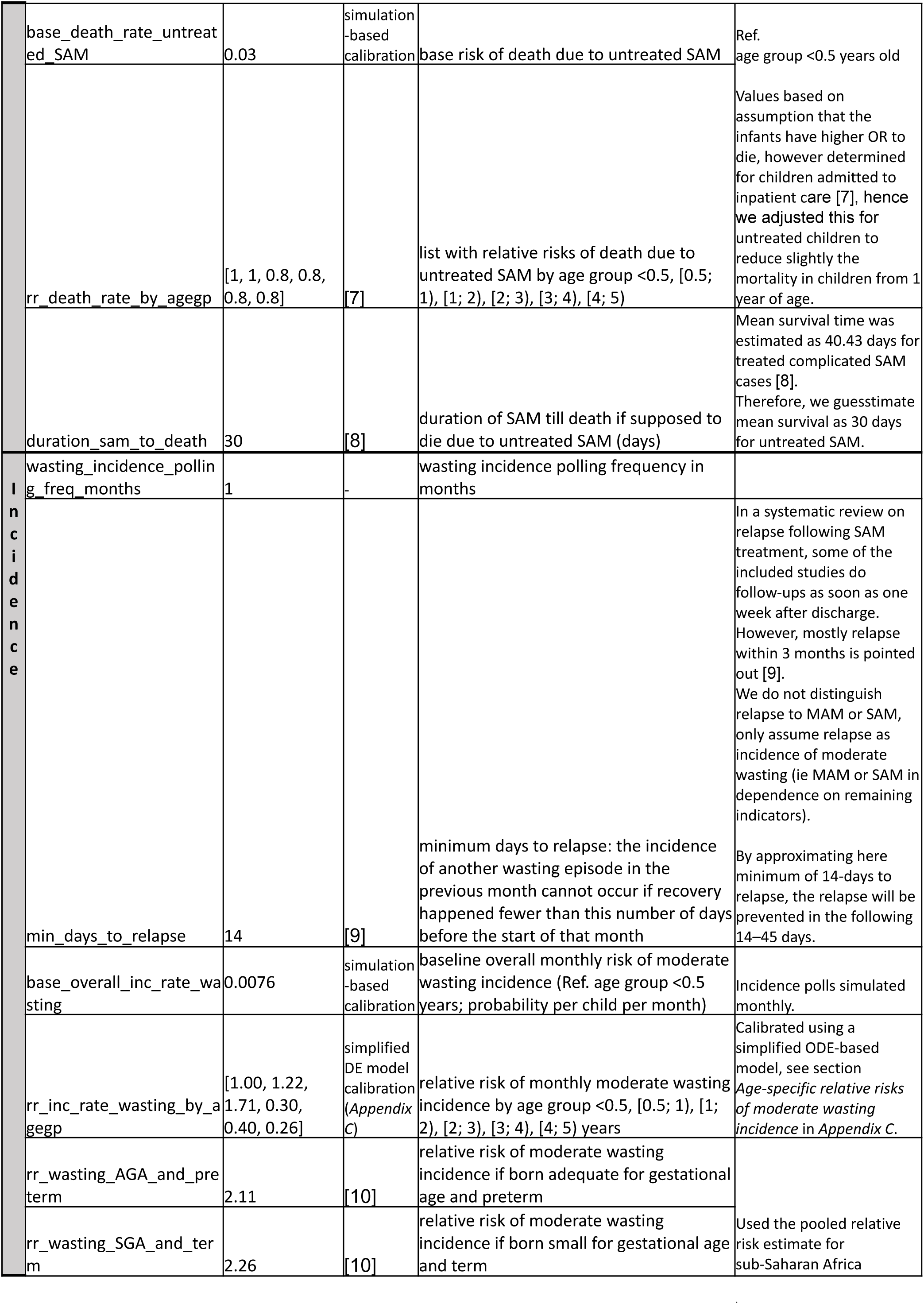
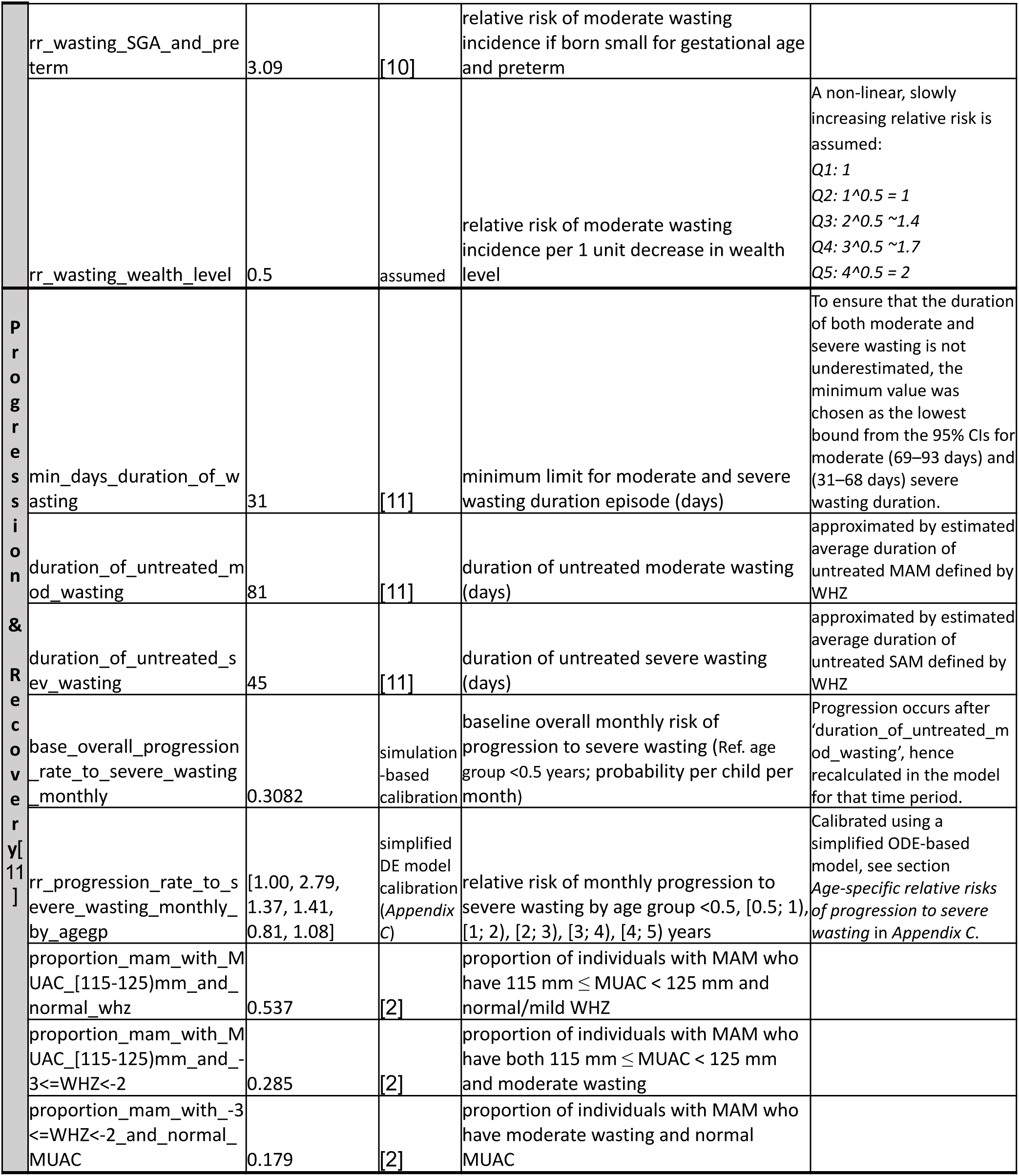
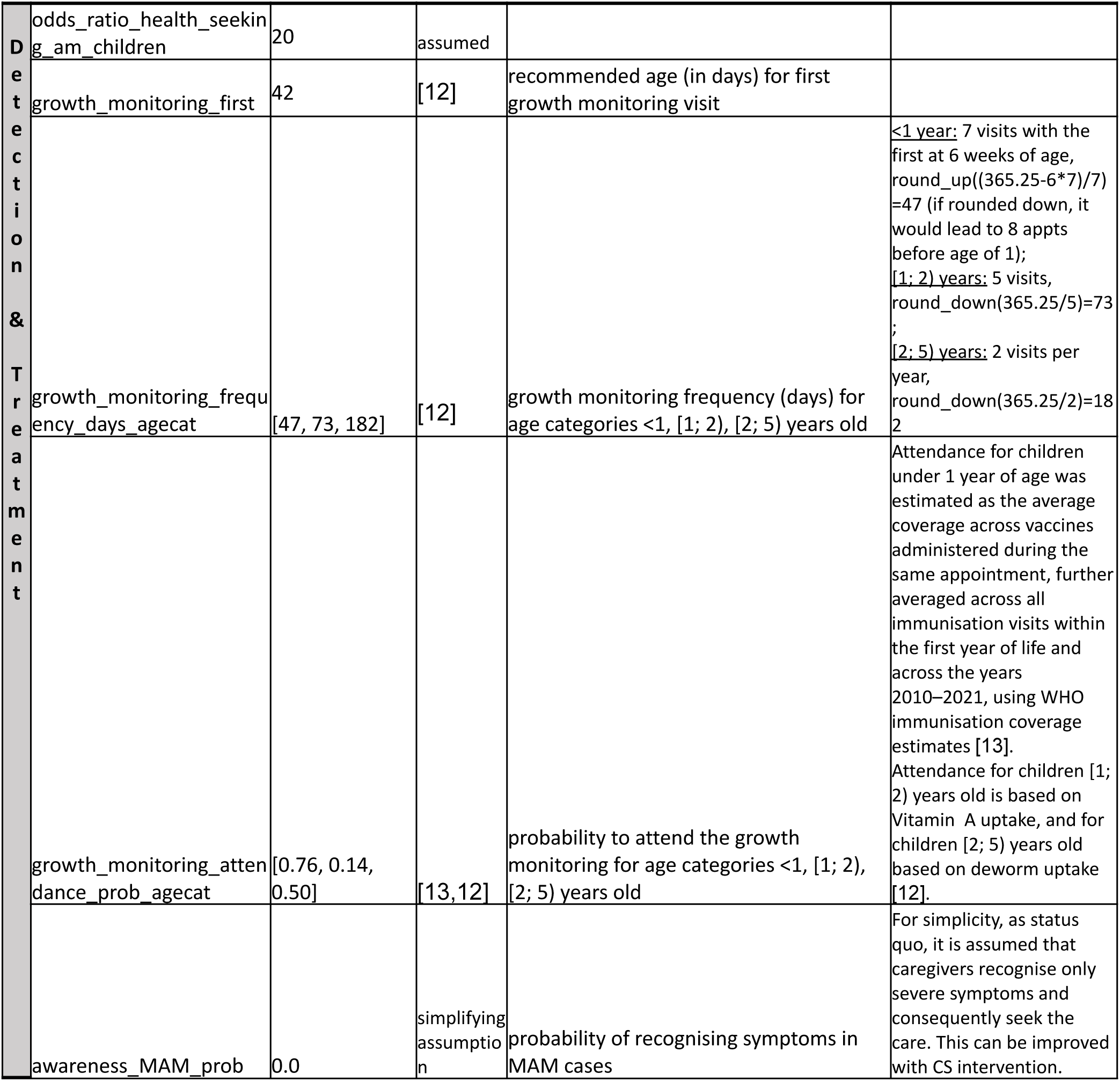
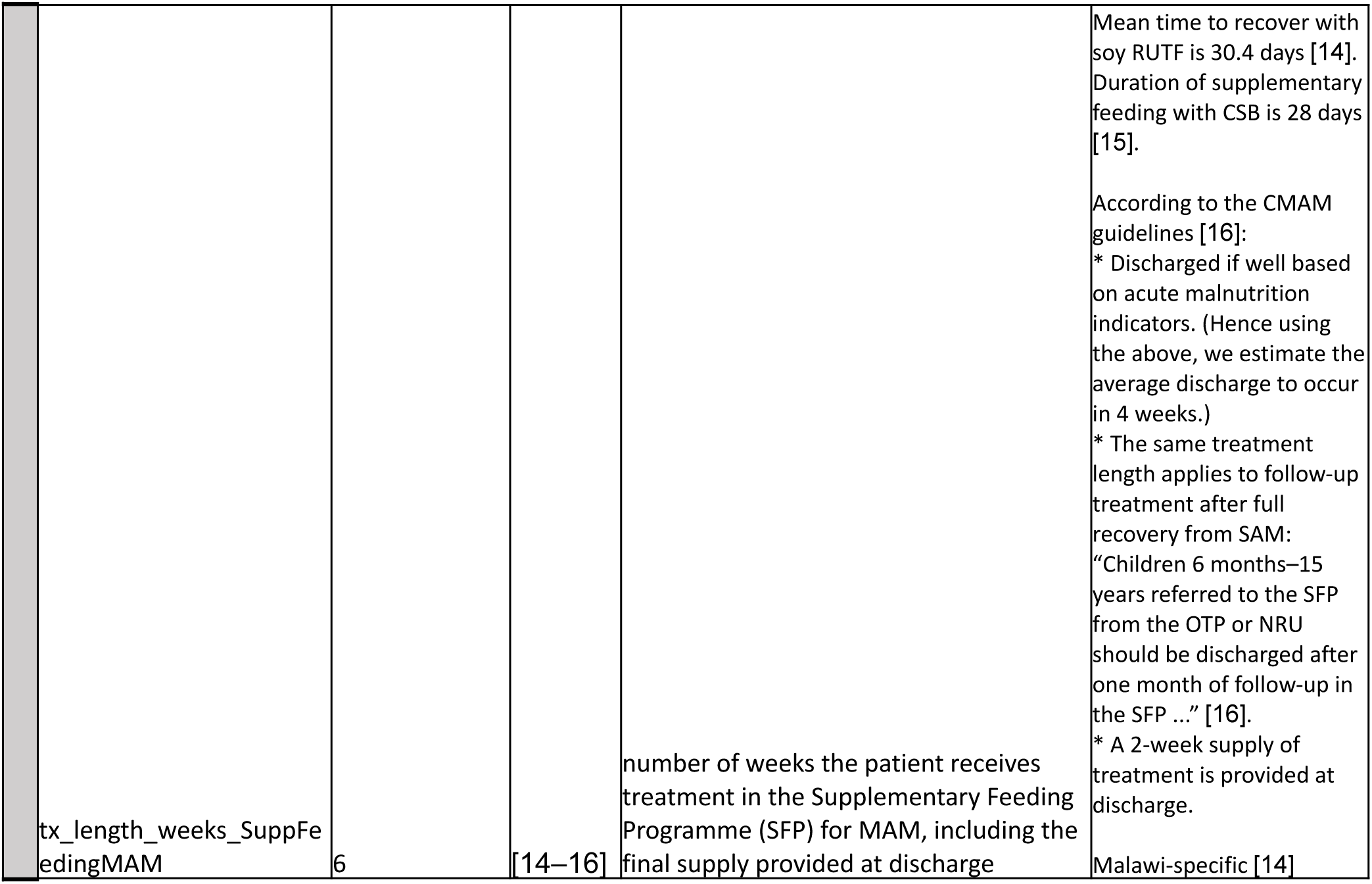
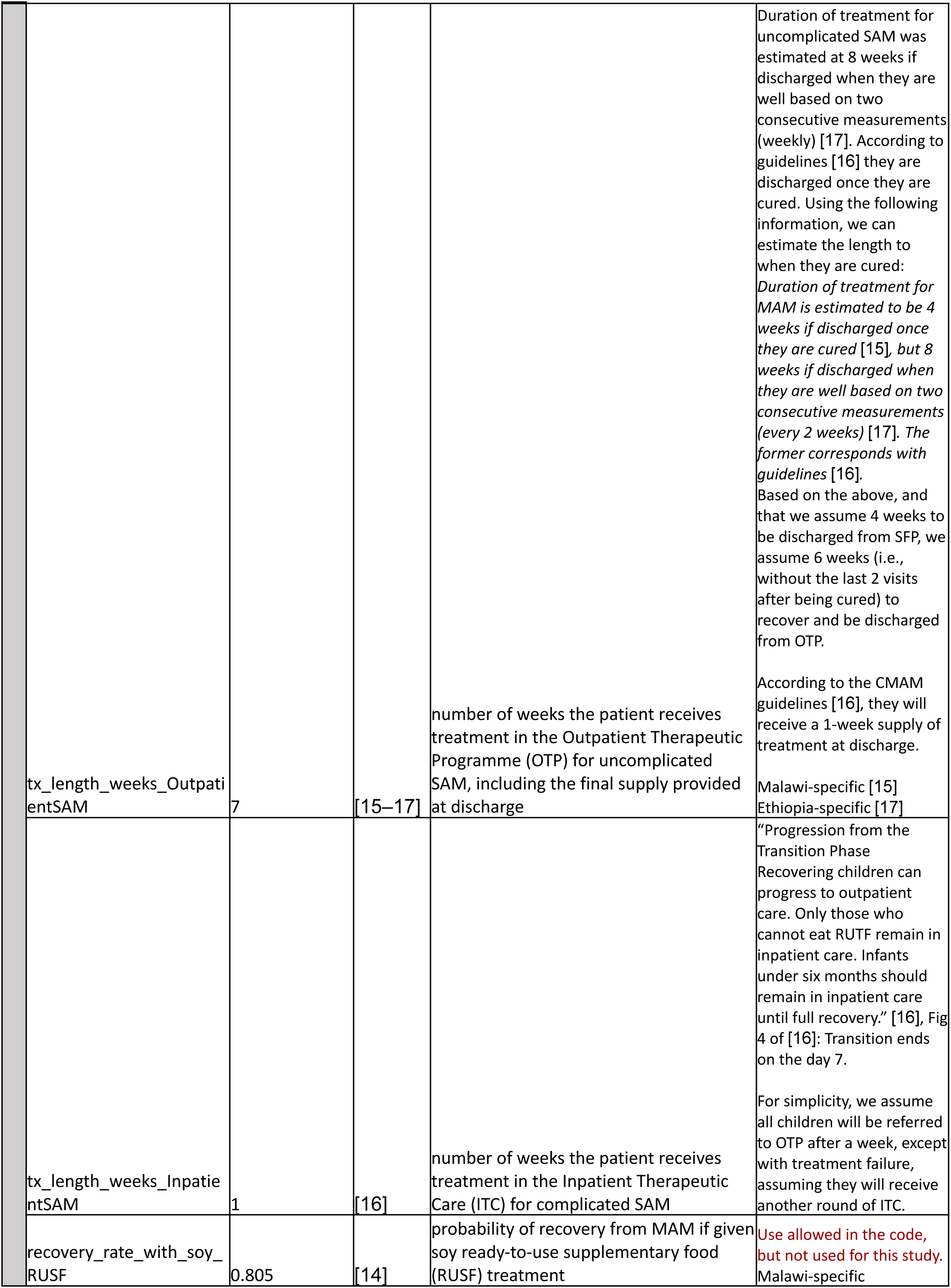
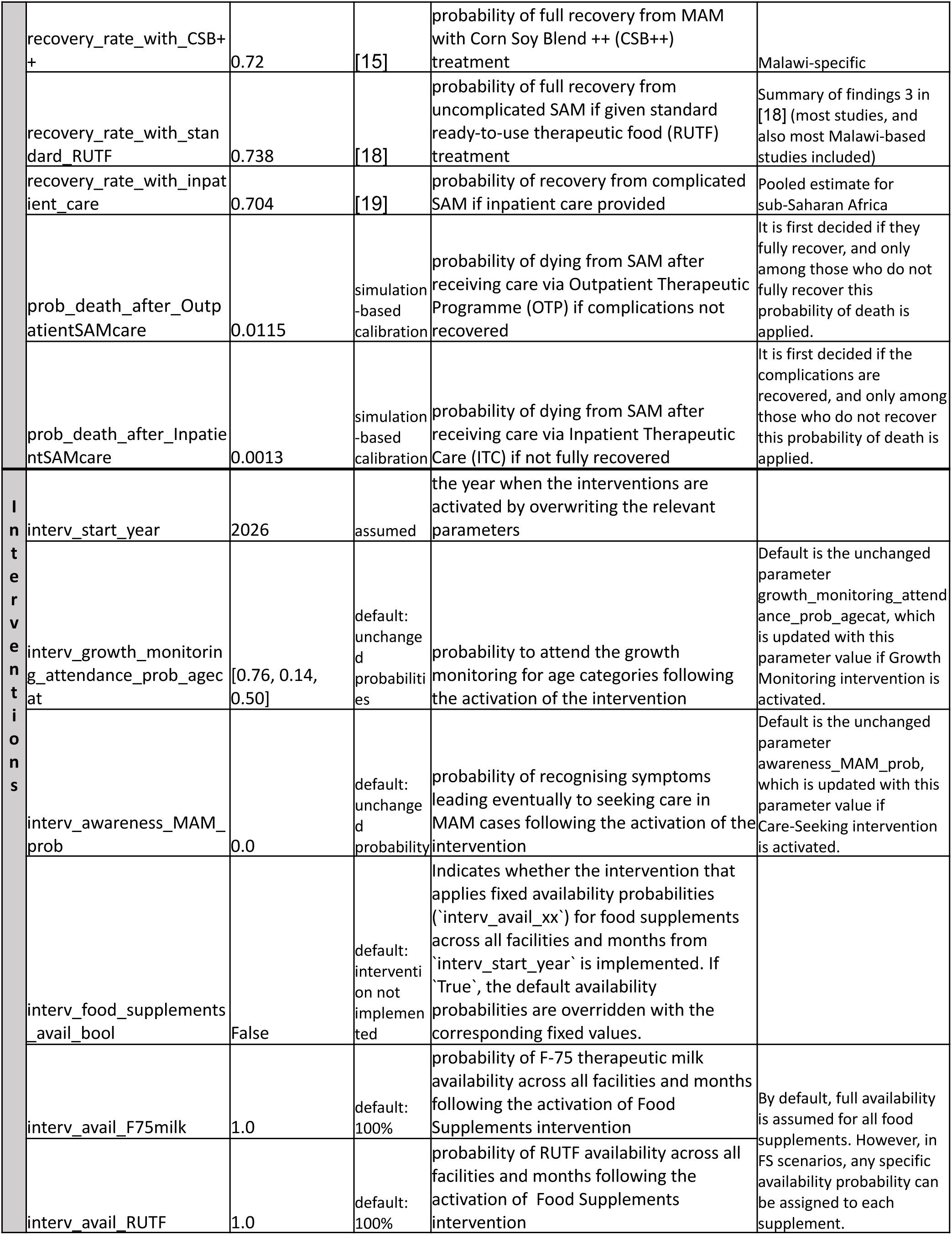
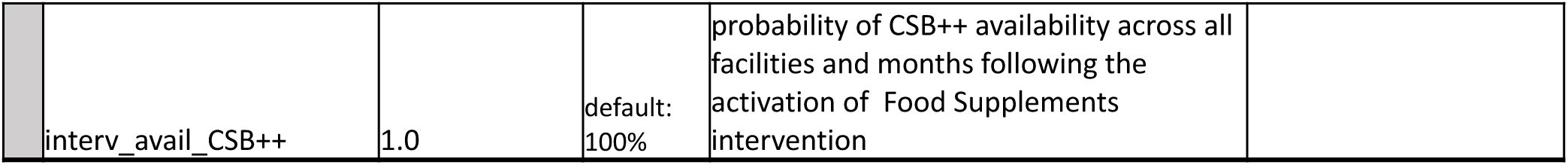
Wasting model parameters.

### Appendix B

#### Detailed model parameters use

The parameter values and references are listed in Table A2 in *<u>Appendix A</u>.* Here we describe in detail how these parameters are used in the *Wasting* model.

##### Initial prevalence

The model is designed to focus exclusively on acute malnutrition in the population of children under five (max_age_child_am). When the population is initiated by the TLO framework for simulation, the WHZ category is set for each child under five years of age. The modelled initial prevalence was informed by DHS 2010 data [1]. The prevalence of any wasting (WHZ < -2) for each age group: 0–5, 6–11, 12–23, 24–35, 36–47, and 48–59 months (prev_init_any_by_agegp), is used to determine whether the child is wasted. The severity of wasting (moderate -3 ≥ WHZ < -2, or severe WHZ < -3) is assigned based on the probability of severe wasting among all wasting cases given by age group (equation (1)), based on DHS 2010 [1] wasting (P_agegp_(WHZ<-2): prev_init_any_by_agegp, and severe wasting prevalence (P_agegp_(WHZ<-3): prev_init_sev_by_agegp).

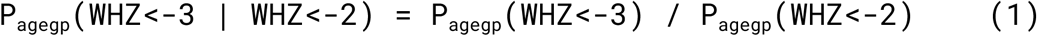

Depending on the assigned WHZ category, including children with normal WHZ (WHZ ≥ -2) assigned, the proportions and distribution of MUAC categories (below 115 mm; 115 mm–125 mm, over 125 mm) and the proportions of oedematous cases are applied to all children under five. It is assumed that children with severe wasting consistently have a MUAC below the normal threshold (i.e., MUAC < 125 mm). In contrast, children younger than six months are simulated to always have a normal MUAC (i.e., MUAC ≥ 125 mm), reflecting the fact that MUAC is typically not measured at this age. This approach ensures that the diagnosis of acute malnutrition in this age group is based solely on the presence of wasting and/or nutritional oedema, see diagnostics criteria in Table 1 in *Methods* section. <u>Table B1</u> shows the parameters used to generate the probabilities for determining MUAC category and oedema presence status, based on the already assigned WHZ category.

**Table B1.**
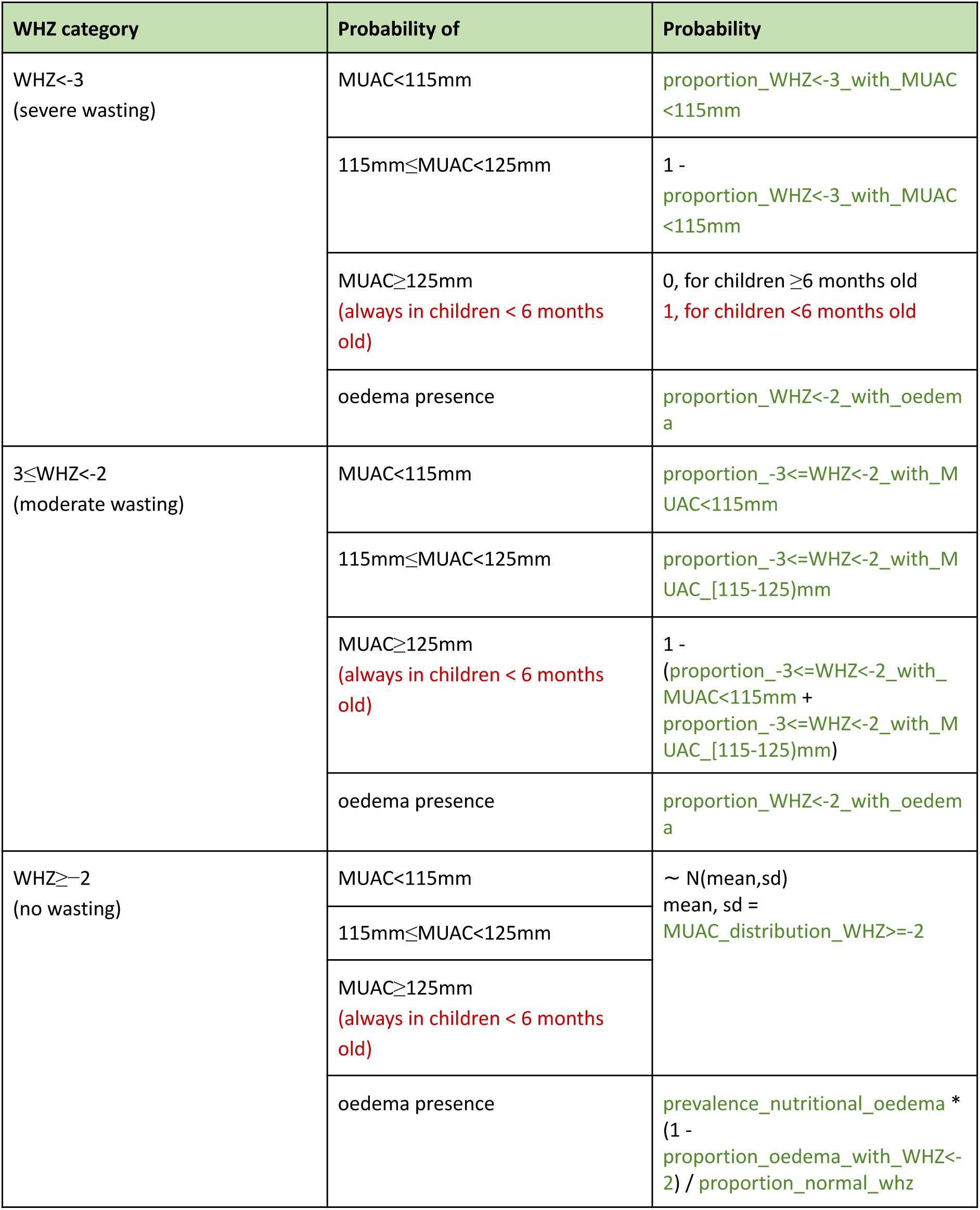
Probabilities of MUAC and oedema presence indicators in dependence of assigned WHZ category.

These three indicators collectively determine the child’s acute malnutrition state according to the criteria from Table 1. For SAM cases presence of complications is determined (prob_complications_in_SAM), and it is determined whether the child will die due to SAM if it remains untreated. The risk of death is assumed to be higher in children less under one year of age (base_death_rate_untreated_SAM, rr_death_rate_by_agegp). If the child is determined to die due to SAM, the death is modelled to occur in 30 days from SAM onset (duration_sam_to_death).

##### Incidence of moderate wasting

The incidence of new wasting cases is determined by several risk factors, including age, size at birth, preterm birth, and household wealth (base_overall_inc_rate_wasting, rr_inc_rate_wasting_by_agegp, rr_wasting_AGA_and_preterm, rr_wasting_SGA_and_term, rr_wasting_SGA_and_preterm, rr_wasting_wealth_level) using linear regression model described in *<u>Appendix C</u>*, which is applied monthly (wasting_incidence_polling_freq_months). Wasting can occur in any child under five who is well-nourished, not currently receiving treatment for acute malnutrition (including follow-up care after recovery from SAM), and has not recovered from acute malnutrition within the 14 days (min_days_to_relapse) prior to the month of incidence onset. This condition is included to prevent premature relapses, although relapses themselves are not explicitly modelled. The modelled annual incidence by age over time is illustrated in *<u>Appendix C</u>* (Fig C1). The remaining indicators (MUAC, and oedema presence) are determined using the probabilities from <u>Table B1</u> for moderate wasting, and acute malnutrition status updated accordingly. In case of onset of SAM, presence of complications (prob_complications_in_SAM) and whether the child will die due to SAM if remains untreated (base_death_rate_untreated_SAM, rr_death_rate_by_agegp), is determined. The incidence is illustrated in the progression part of <u>Fig B1</u> (no wasting -> moderate wasting).

**Fig B1.**
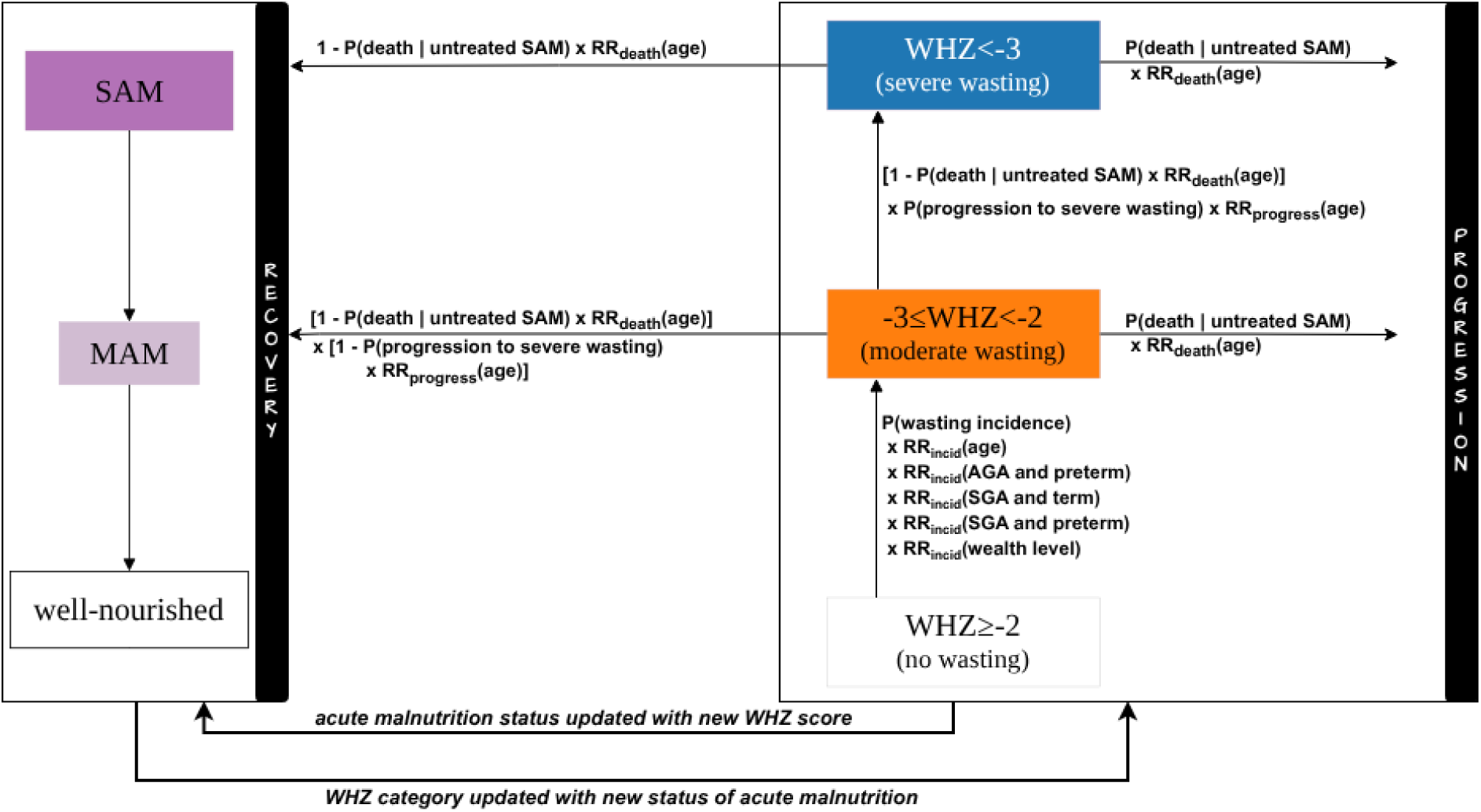
Detailed modelled natural history without treatment.

##### Progression & Recovery

The modelled natural history of wasting progression and recovery in the absence of interaction with the healthcare system is outlined in <u>Fig B1</u>, representing how the condition evolves when individuals do not receive any care.

After each change, due to incidence, progression or recovery, if the child is moderately wasted, the episode lasts 81 days (duration_of_untreated_mod_wasting), if the child is severely wasted, the episode lasts 45 days (duration_of_untreated_sev_wasting), after this time the child may again progress or recover [11], however if they are determined to die due to SAM, the death occurs 30 days (duration_sam_to_death) from the onset of SAM [8].

The base monthly risk of natural progression to severe wasting depends only on age (base_overall_progression_rate_to_severe_wasting_monthly, rr_progression_rate_to_severe_wasting_monthly_by_agegp), and the progression is modelled through the severity of wasting (moderate wasting -> severe wasting or moderate wasting -> death, and severe wasting -> death), as shown in the progression part of <u>Fig B1</u>. Since the progression to severe wasting occurs after the duration of untreated severe wasting of 45 days, the monthly risks are consequently recalculated for this time period for the use within the model. The remaining indicators (MUAC, and oedema presence) are determined using the probabilities from <u>Table B1</u> for severe wasting.

On the contrary, natural recovery is based on the acute malnutrition state (SAM -> MAM -> well-nourished) regardless of the severity of wasting, as shown in the recovery part of <u>Fig B1</u>. When a child recovers from SAM without complications to MAM (SAM->MAM recovery), the WHZ and MUAC are assigned according to the proportions of these indicators among MAM cases (proportion_mam_with_MUAC_[115-125)mm_and_normal_whz, proportion_mam_with_MUAC_[115-125)mm_and_-3<=WHZ<-2, proportion_mam_with_-3<=WHZ<-2_and_normal_MUAC). With full recovery, all indicators and the malnutrition state are set to correspond with being well-nourished (MAM->well-nourished recovery).

##### Health System Interactions

The CMAM programme [16] implements four components, which are all modelled:

- community outreach and facility growth monitoring activities,
- SFP for MAM,
- OTP for SAM without medical complications, and
- ITC for SAM with medical complications.

The growth monitoring, SFP, and OTP are delivered at primary care facilities (primary level), while the ITC at NRU or in the children’s ward at a health facility with 24-hour care (secondary level) [20].

###### Detection

The detection of acute malnutrition can happen in two ways, via healthcare seeking, or during a routine growth monitoring.

###### Care-seeking

Severely malnourished children often develop severe symptoms, which are very likely to prompt caregivers to seek medical attention. In contrast, moderate malnutrition is frequently unrecognised or not perceived as serious [21,22]. The model assumes 100% awareness of presence of acute malnutrition symptoms in cases of SAM, while we assume no awareness when the child is only moderately malnourished (awareness_MAM_prob). The odds of seeking care (for any reason) are assumed to be 20 times greater if caregivers are aware of symptoms of acute malnutrition in the child (odds_ratio_health_seeking_am_children), meaning that over 90% of children with SAM are seeking care. Although MAM does not lead to seeking care by itself, malnutrition is a component of the IMCI [23] and, consequently, if children seek medical care for other symptoms—whether related or unrelated to malnutrition—they will be examined for malnutrition as part of the routine assessment in non-emergency cases, when both MAM and SAM can be detected. The care can be provided at any facility level.

###### Routine growth monitoring

In the model, children are born well-nourished regardless of preterm or low birth weight status. During the first year of life, growth monitoring, screening for both MAM and SAM, is conducted through the Expanded Programme on Immunisation, which includes 7 immunisation appointments with the first growth monitoring recommended at 6 weeks of age (growth_monitoring_first). Between the ages of 1 and 2 years, children are recommended to attend 5 visits for growth monitoring, and from 2 to 5 years of age, children are recommended to receive growth monitoring, every 6 months [12]. Appropriate to the number of recommended growth monitoring appointments per year, we calculated the number of days between two appointments for each age category (growth_monitoring_frequency_days_agecat). Based on WHO immunisation coverage estimates, we estimated the average probability to attend each immunisation appointment for children younger than one year to be 76% [13].

However, the attendance drops once not integrated with immunisation, to 14% for children of age 1–2 years, and to 50% for children of age 2–5 years as estimated in [12] (growth_monitoring_attendance_prob_agecat). On the day when the child is supposed to attend an appointment for growth monitoring, based on the attendance probability it is determined whether the child does or does not attend this particular appointment. If it does, the assessment of acute malnutrition is carried out.

###### Treatment

If acute malnutrition is detected, following the Malawi CMAM guidelines [16], the child is referred to a corresponding treatment programme: SFP for MAM cases, OTP for SAM cases without medical complications, and ITC for SAM cases with medical complications. Admission to the treatment is conditional to availability of corresponding food supplement, if not available the child will not be admitted for the treatment, however, in future can be due to detection via care-seeking or growth monitoring referred for the treatment again.

The assessment for acute malnutrition at the point of enrolment into treatment, including follow-up, is simulated. However, ongoing assessments conducted during treatment (bi-weekly during SFP, weekly during OTP, and daily during ITC) are not simulated, and treatment delivery is modelled as a consolidated event, with consumables for the entire course of treatment recorded as being administered at the initiation of treatment, and outcomes happening at the end of the treatment.

Note that, although SAM may result in death during treatment, treatment can fail and child be consequently referred for a follow-up treatment, the severity of acute malnutrition—whether MAM or uncomplicated SAM—is assumed to not worsen otherwise while the child is receiving care, see <u>Fig B2</u>.

**Fig B2.**
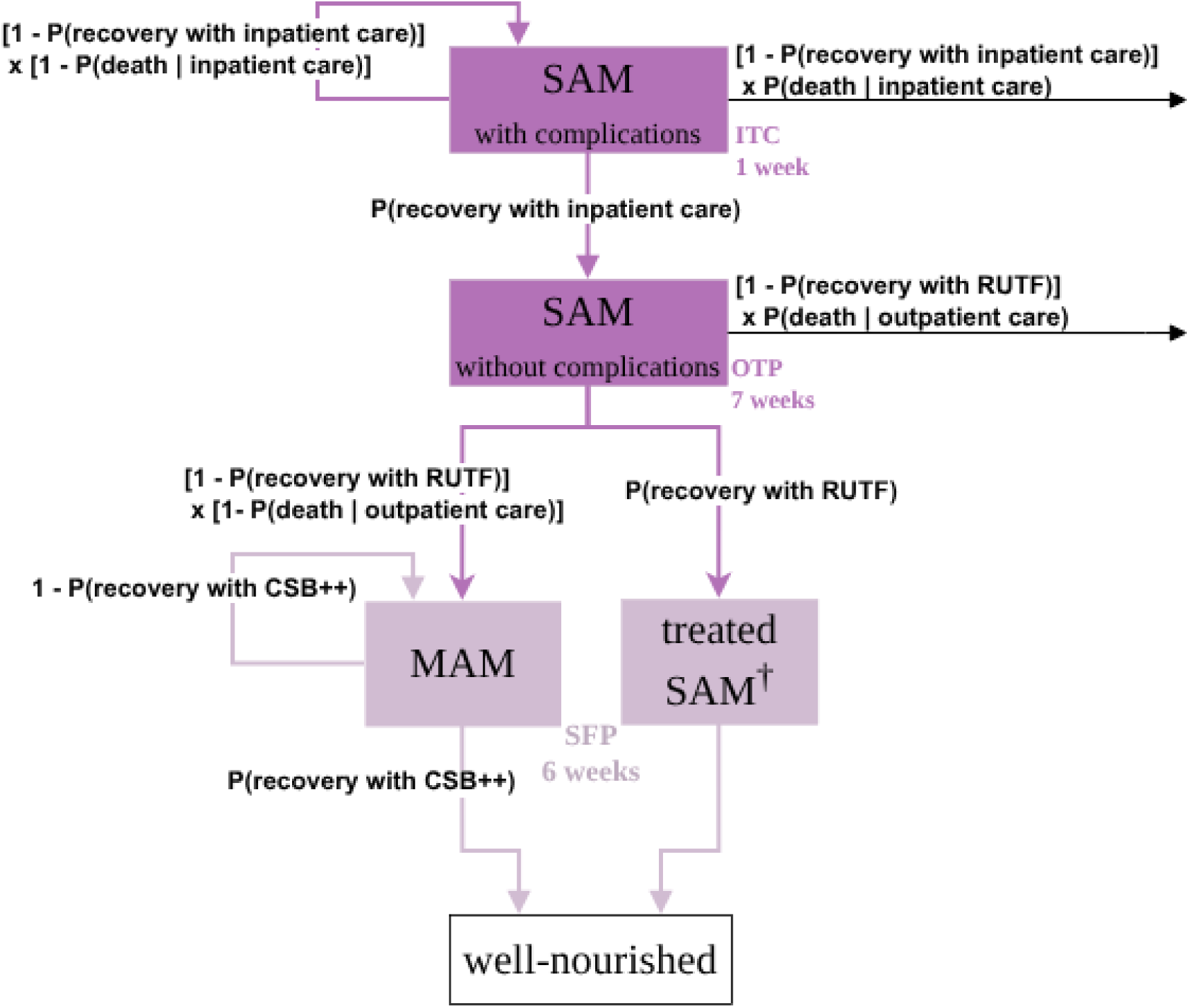
Detailed modelled treatment outcomes. ^†^treated SAM, refers to children who have fully recovered from SAM and require follow-up supplementary feeding through a SFP

###### Supplementary feeding programme (SFP)

Supplementary feeding programme (SFP) targets children (0–15 years), and pregnant and lactating women up to six months postpartum, however, the *Wasting* model only focuses on under-five children. The programme is delivered to children with MAM, or those discharged from OTP after full recovery from SAM, who are referred to SFP for continuum of care to prevent a re-occurrence of malnutrition.

The nutritional treatment is based on CSB++, alternatively Soy Ready-to-Use Supplementary Food (RUSF) could be used, but according to current guidelines only CSB++ (for under-5 years old) and CSB+ (for 5–15 years old, pregnant or lactating women) are administered. Accordingly, the CSB++ is administered by the *Wasting* model as a type of SFP, the option to switch the treatment to the alternative RUSF treatment is available if future studies require.

The supplies of CSB++ are administered every 2 weeks along with measurements of the malnutrition indicators being performed (except height or length measurement, that is performed monthly) for 4 weeks, and a final 2 weeks’ supply administered at discharge. The model simplifies the process by simulating the entire treatment duration as the time required to consume all food supplements, consequently, the treatment is modelled to last 6 weeks (tx_length_weeks_SuppFeedingMAM). The child can either fully recover (recovery_rate_with_CSB++) with treatment, in such cases the acute malnutrition state and the indicators are updated from ‘MAM’ to ‘well-nourished’ and the child is discharged from the treatment, or, when treatment is not successful, remain MAM and is referred for another SFP cycle.

Children are also referred for follow-up SFP after full recovery from SAM (when their acute malnutrition status is ‘well-nourished’) and, subject to availability, receive CSB++ for 6 weeks before being discharged from treatment.

###### Therapeutic Programme (OTP)

OTP provides home-based treatment with RUTF for children with SAM who have no medical complications, modelled simply as presence or absence of complications.

Medical treatment in OTP includes antibiotic therapy with amoxicillin, malaria testing for all children using a rapid diagnostic test (mRDT) followed by appropriate treatment, deworming with albendazole or mebendazole, and administration of measles vaccine if the child has not been previously vaccinated. The tests are not currently explicitly modelled as part of OTP, the medicine is modelled as a package administered if available but not essential for admission to the treatment. The RUTF is administered weekly for 6 weeks with 1-week supply at discharge, which is in model simplified as 7 weeks long treatment (tx_length_weeks_OutpatientSAM). The child can fully recover (recovery_rate_with_standard_RUTF) with treatment as well-nourished, however if the treatment is not successful, the child may either die due to SAM (prob_death_after_OutpatientSAMcare), or recover to MAM. When recovered to MAM or fully as well-nourished, the child is referred to a follow-up SFP, to complete the recovery or prevent relapse, respectively.

###### Inpatient therapeutic care (ITC)

Inpatient therapeutic care targets children with SAM up to 15 years old who present with any of the danger signs or medical complications, although the *Wasting* model focuses exclusively on children under five. Also, in reality, children under six months of age with SAM or feeding difficulties who are losing or not gaining weight would be provided ITC even if no complications are present, however, the model does not specifically simulate the weight, hence this aspect is not reflected. The danger signs include anorexia, no appetite, intractable vomiting, convulsions, lethargy, not being alert, unconsciousness, inability to drink or breastfeed, and fever (> 39° C rectal or > 38.5° C axillary). The medical complications include severe anaemia, cardiac failure, severe dermatosis, signs of vitamin A deficiency, diarrhoea with dehydration, and severe malaria. These are modelled simply as presence of complications in SAM cases.

Medical treatment in ITC includes antibiotics, measles vaccine if not vaccinated, Anti-Malarial if tested positive, glucose if low or not able to measure it, oral rehydration solution (ORS) ReSoMal if dehydrated or having watery stool, antiretroviral therapy (ART) for HIV positive, vitamin A with active measles or vitamin A deficiency. The tests are not explicitly modelled, instead, the medicine is modelled to be administered as a package if available but not essential for admission to the treatment. F-75 therapeutic milk is administered for the first three days (stabilisation phase), followed by four days of RUTF (transition phase), both food supplements mandatory for admission as previously defined. Care and food supplements are provided daily, with feeds given every few hours during both the stabilisation and transition phases. At the end of the treatment period, after 1 week (tx_length_weeks_InpatientSAM), if the treatment is successful (recovery_rate_with_inpatient_care), the complications are resolved and the child is referred to OTP. If the treatment is unsuccessful, either the child dies due to SAM (prob_death_after_InpatientSAMcare), or the complications persist and the child is referred for another week of ITC.

###### Consumables

The consumables required for the treatments, the average amount being dispensed per case if available, and its cost are shown in <u>Table B2</u>. The seasonal average availability probabilities of essential consumables in the facility level where the treatment is provided, primary level for SFP and OTP, and secondary level for ITC, for each month in a year are presented in <u>Fig B3</u>, and are translated to average probabilities of treatments being available, shown in Fig 2 in *Methods* section. There are more stock outs in the first months of the year, which is consistent with higher prevalence of wasting in the rainy season particularly between January and March [24], however we do not explore this any further in our study.

**Table B2.** Treatment Consumables.

| Treatment | Item | Essential/Non-essential for treatment admission | Total dispensed amount per case [16] | Case cost 2023 USD |
| --- | --- | --- | --- | --- |
| SFP | Corn Soya Blend (or Supercereal - CSB++)[25] | essential | 9,000 g | 43 [25] |
| OTP | RUTF | essential | 140 of 92g sachets | 45 [25] |
|  | SAM medicines | non-essential | 1 package<sup>§</sup> | 2 [26] |
| ITC | F-75 milk (stabilisation phase) | essential | 2,460 ml<sup>#</sup> | 4 [25] |
|  | RUTF (transition phase) | essential | 12 of 92g sachets | 1 [25] |
|  | SAM medicines | non-essential | 1 package<sup>‡</sup> | 2 [26] |

SFP = Supplementary Feeding Programme for MAM
OTP = Outpatient Therapeutic Programme for SAM with no complications
ITC = intensive Inpatient Therapeutic Care for SAM with complications
<sup>§</sup> antibiotics with Amoxicillin supply for 7 days at admission, Albendazole or Mebendazole 1st dose 7 days after admission and 2nd dose after 6 months, measles vaccine at admission if not vaccinated or if ongoing epidemic, Anti-Malarial if tested positive [16]
<sup>#</sup> 90 ml per feed for children with oedema<sup>†</sup> and 115 ml for children with severe wasting<sup>†</sup>, averaged to 102.5 ml per feed [16]
<sup>†</sup> bodyweight of 7 kg is used for estimating costs as recommended [27]
<sup>‡</sup> broad spectrum IV antibiotics for 7 days, Albendazole or Mebendazole 1st dose 7 days after admission and 2nd dose after 6 months, measles vaccine at admission if not vaccinated, Anti-Malarial if tested positive, glucose if low or not able to measure it, oral rehydration solution (ORS) ReSoMal if dehydrated or watery stool, ART for HIV positive, vitamin A with active measles or vitamin A deficiency [16]

**Fig B3.**
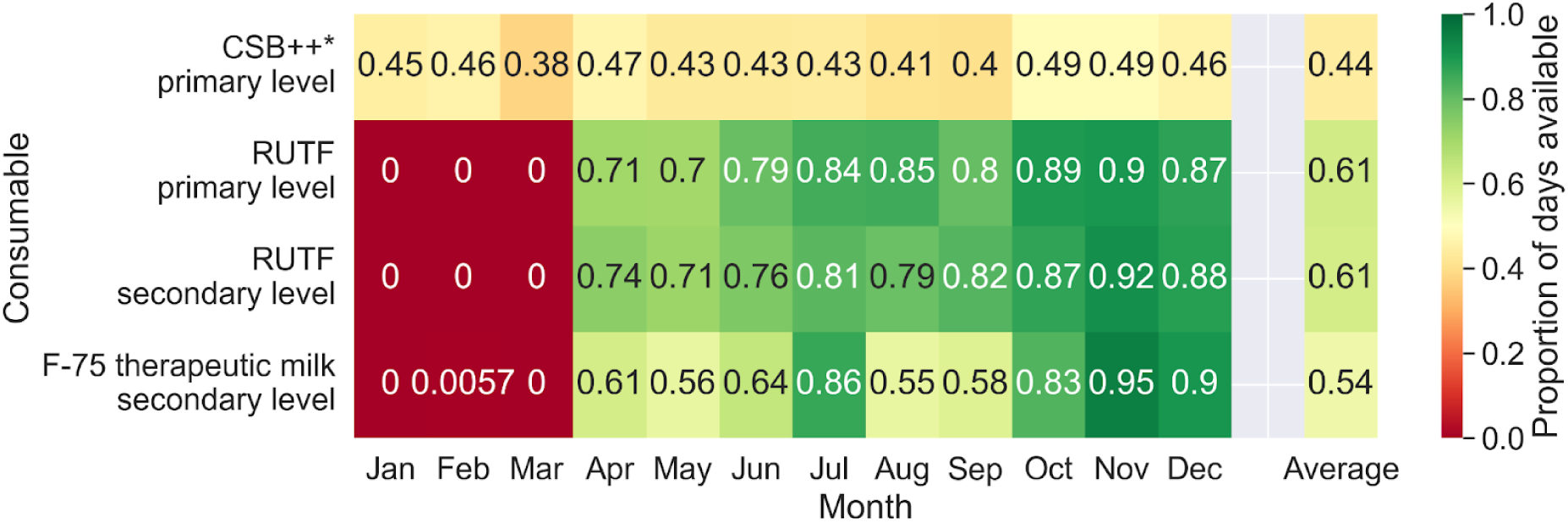
Monthly average availability of food supplements at corresponding facility level. *The data on CSB++ were not feasible, therefore, their availability was approximated by average availability across all consumables at the facility level with feasible availability data.

#### Calibration

Model calibration is described in the *Methods* section. Here are presented the final calibrated values for the risks (<u>Table B3</u>), which were then translated into the parameters:

- baseline risks and age-specific relative risks of monthly moderate wasting incidence (base_overall_inc_rate_wasting, rr_inc_rate_wasting_by_agegp,) and monthly progression to severe wasting (base_overall_progression_rate_to_severe_wasting_monthly, rr_progression_rate_to_severe_wasting_monthly_by_agegp),
- baseline risks of death due to untreated SAM (base_death_rate_untreated_SAM),
- death due to SAM when treated with outpatient (prob_death_after_OutpatientSAMcare) or inpatient care (prob_death_after_InpatientSAMcare).

Their values are listed in Table A2 in *<u>Appendix A</u>*.

**Table B3.** Calibrated values.

| Parameter | Calibrated value by age group |  |  |  |  |  |
| --- | --- | --- | --- | --- | --- | --- |
|  | 0–5 mo | 6–11 mo | 12–23 mo | 24–35 mo | 36–47 mo | 48–59 mo |
| monthly risk of moderate wasting incidence | 0.76% | 0.93% | 1.30% | 0.23% | 0.30% | 0.20% |
| monthly risk of progression to severe wasting | 30.82% | 86.14% | 42.29% | 43.37% | 25.08% | 33.21% |
| mortality risk among untreated SAM cases | 3.00% | 3.00% | 2.40% | 2.40% | 2.40% | 2.40% |
| mortality risk among unsuccessfully treated SAM cases* | 0.30% | 0.30% | 0.30% | 0.30% | 0.30% | 0.30% |

mo = months
\* unsuccessfully treated SAM cases refer to patients who do not reach full recovery during OTP or whose complications are not resolved during ITC

#### Scenarios

Theoretical maximum scenarios are simulated, assuming the constraints if addressed are fully eliminated (full attendance to all recommended growth monitoring appointments in children 1–5 years; awareness of symptoms in 100% MAM cases; and/or all food supplements for acute malnutrition treatment always available in requisite amounts), although not realistic, allows to evaluate potential impact of the constraints being addressed. Each scenario adopts the same conditions as the status quo (SQ) prior to 2026, with scenario-specific changes simulated from the first day of 2026 onwards. The values related to constraints considered for each scenario are summarised in <u>Table B4</u>.

**Table B4.**
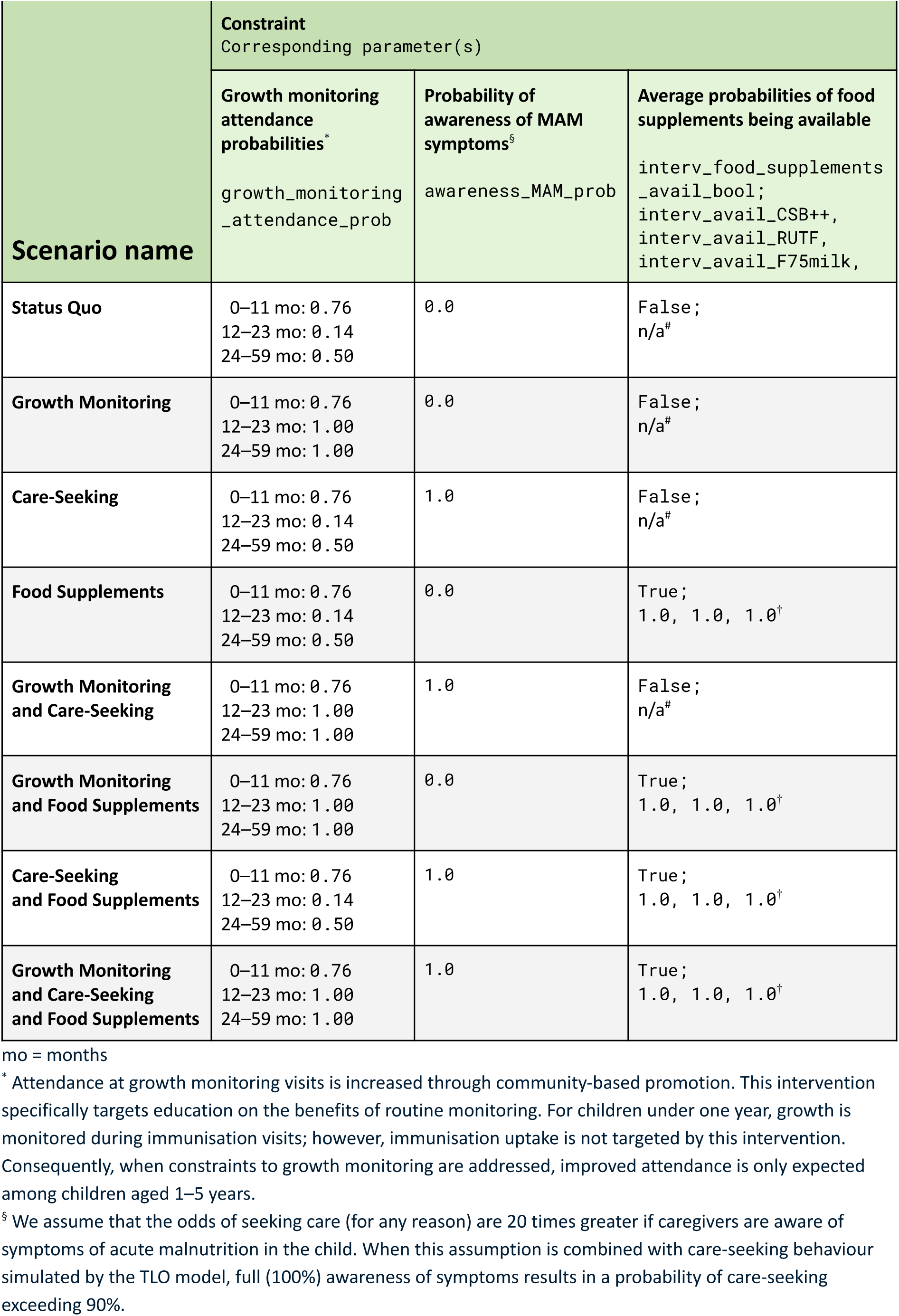
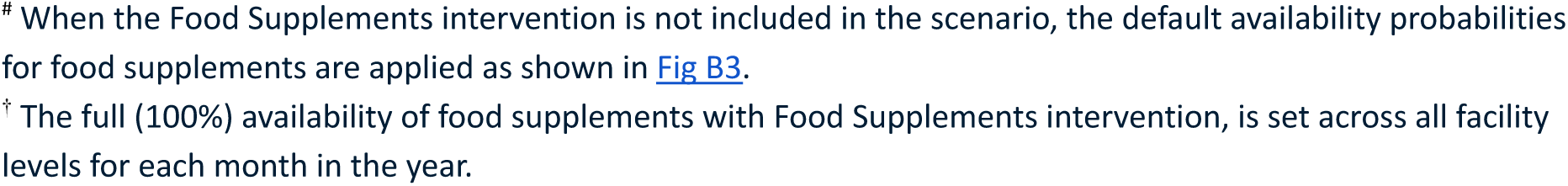
Scenarios addressing constraints in acute malnutrition management in under-five children.

| Scenario name | Constraint<br>Corresponding parameter(s) |  |  |
| --- | --- | --- | --- |
|  | Growth monitoring attendance probabilities*<br>growth_monitoring_attendance_prob | Probability of awareness of MAM symptoms<sup>§</sup><br>awareness_MAM_prob | Average probabilities of food supplements being available<br>interv_food_supplements_avail_bool;<br>interv_avail_CSB++,<br>interv_avail_RUTF,<br>interv_avail_F75milk, |
| Status Quo | 0–11 mo: 0.76<br>12–23 mo: 0.14<br>24–59 mo: 0.50 | 0.0 | False;<br>n/a<sup>#</sup> |
| Growth Monitoring | 0–11 mo: 0.76<br>12–23 mo: 1.00<br>24–59 mo: 1.00 | 0.0 | False;<br>n/a<sup>#</sup> |
| Care-Seeking | 0–11 mo: 0.76<br>12–23 mo: 0.14<br>24–59 mo: 0.50 | 1.0 | False;<br>n/a<sup>#</sup> |
| Food Supplements | 0–11 mo: 0.76<br>12–23 mo: 0.14<br>24–59 mo: 0.50 | 0.0 | True;<br>1.0, 1.0, 1.0<sup>†</sup> |
| Growth Monitoring and Care-Seeking | 0–11 mo: 0.76<br>12–23 mo: 1.00<br>24–59 mo: 1.00 | 1.0 | False;<br>n/a<sup>#</sup> |
| Growth Monitoring and Food Supplements | 0–11 mo: 0.76<br>12–23 mo: 1.00<br>24–59 mo: 1.00 | 0.0 | True;<br>1.0, 1.0, 1.0<sup>†</sup> |
| Care-Seeking and Food Supplements | 0–11 mo: 0.76<br>12–23 mo: 0.14<br>24–59 mo: 0.50 | 1.0 | True;<br>1.0, 1.0, 1.0<sup>†</sup> |
| Growth Monitoring and Care-Seeking and Food Supplements | 0–11 mo: 0.76<br>12–23 mo: 1.00<br>24–59 mo: 1.00 | 1.0 | True;<br>1.0, 1.0, 1.0<sup>†</sup> |

mo = months
\* Attendance at growth monitoring visits is increased through community-based promotion. This intervention specifically targets education on the benefits of routine monitoring. For children under one year, growth is monitored during immunisation visits; however, immunisation uptake is not targeted by this intervention. Consequently, when constraints to growth monitoring are addressed, improved attendance is only expected among children aged 1–5 years.
<sup>§</sup> We assume that the odds of seeking care (for any reason) are 20 times greater if caregivers are aware of symptoms of acute malnutrition in the child. When this assumption is combined with care-seeking behaviour simulated by the TLO model, full (100%) awareness of symptoms results in a probability of care-seeking exceeding 90%.
<sup>#</sup> When the Food Supplements intervention is not included in the scenario, the default availability probabilities for food supplements are applied as shown in Fig B3.
<sup>†</sup> The full (100%) availability of food supplements with Food Supplements intervention, is set across all facility levels for each month in the year.

#### Outcomes

##### Primary outcomes

Analyses focus on overall DALYs. For example, if an intervention prevented a death from severe acute malnutrition (SAM) but the child died within the observed time period from another cause, this was not counted as an averted death. Acute malnutrition is an underlying risk factor for acute lower respiratory infection (ALRI) and diarrhoea (more details in *Methods* section). Accordingly, we also report cause-specific deaths and DALYs attributable to SAM, ALRI, and diarrhoea in <u>Appendix D.</u>

##### DALY weights

DALYs from premature death were defined as the difference between age at death and life expectancy of 90 years as recommended by WHO [28]. Disability from all causes (related and unrelated to acute malnutrition) was measured monthly and combined additively [29]. DALY weights associated with acute malnutrition are shown in <u>Table B5</u>.

**Table B5.**
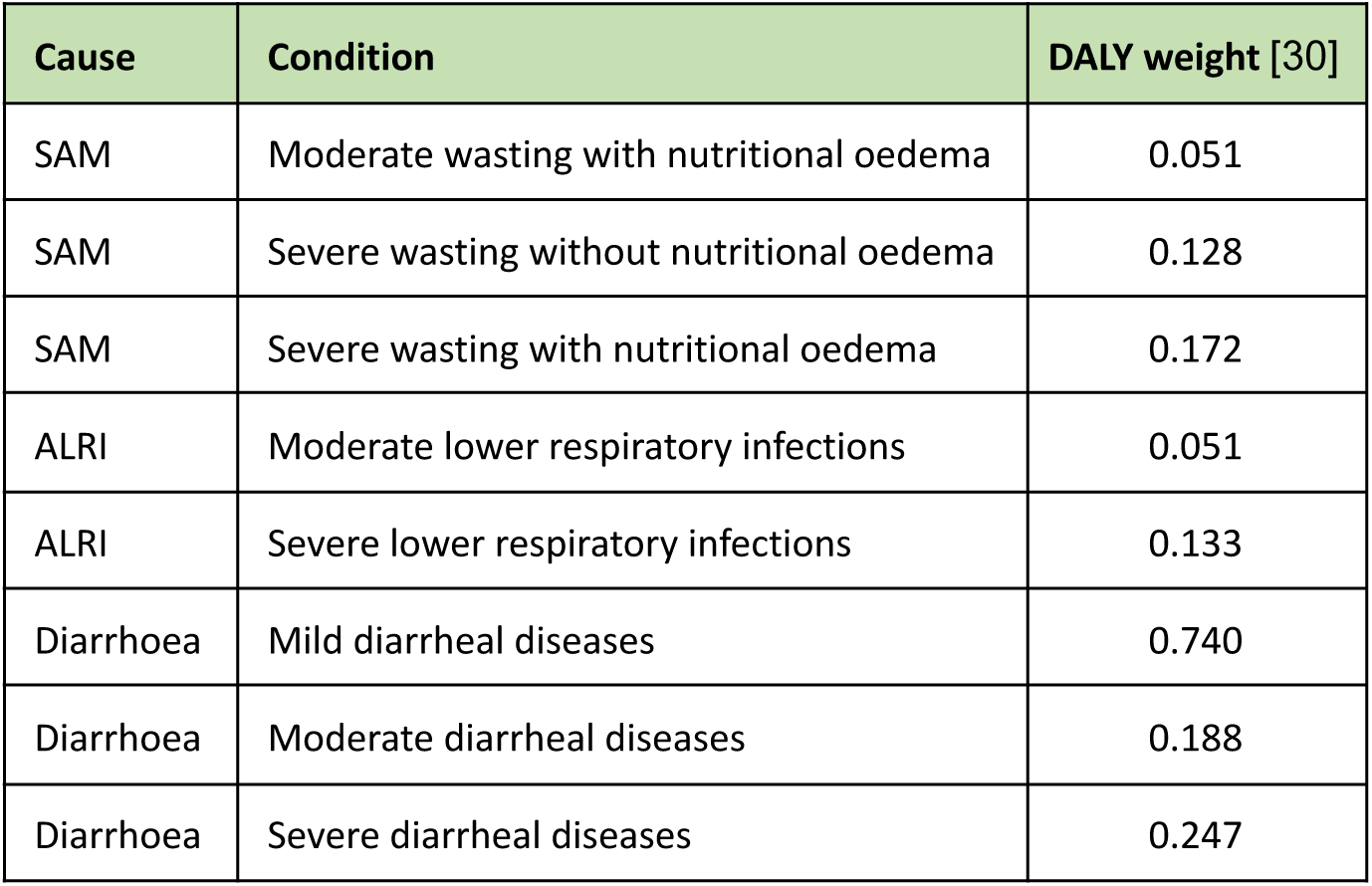
Disability weights associated with acute malnutrition.

##### Costs and Sensitivity analysis

Modelled costs include medical consumables for the treatment of acute malnutrition, as well as allowances for wastage (due to expiry, theft, or mismanagement) and supply chain costs related to procurement, storage, and distribution [31]. No other implementation costs (e.g., health worker training and salaries, communication, administration, travel, preparation, printing, and distribution of materials, and diagnostic tools) are included in the modelled estimates; therefore, we derive separate estimates for additional implementation costs. While total incremental costs are shown in Table 2 in the *Results* section, their distribution between incremental consumable-related costs and additional implementation costs is reported in <u>Table B6</u>, and the disaggregation of the incremental consumable-related costs by cost category is shown in <u>Fig B4</u>.

**Table B6.** Disaggregation and threshold of total incremental costs.

| Scenario | Incremental consumable-related costs vs SQ (2023 USD) | Additional implementation costs (2023 USD) | Maximum allowable total incremental costs (2023 USD, 95% CI) |
| --- | --- | --- | --- |
| *Status Quo* | 0 | 0 | 0 |
| *Care-Seeking* | 2,129,526 | 1,450,643 | 9,413,259<br>(-4,252,392; 23,078,909) |
| *Growth Monitoring* | 3,392,345 | 1,450,538 | 5,774,778<br>(-2,564,620; 14,114,175) |
| *Growth Monitoring and Care-Seeking* | 4,816,380 | 2,175,852 | 17,332,206<br>(5,653,154; 29,011,258) |
| *Care-Seeking and Food Supplements* | 32,331,302 | 1,581,286 | 63,875,745<br>(51,836,347; 75,915,143) |
| *Growth Monitoring and Care-Seeking and Food Supplements* | 33,750,568 | 2,305,639 | 64,332,643<br>(54,925,198; 73,740,088) |
| *Growth Monitoring and Food Supplements* | 35,845,981 | 1,601,635 | 59,716,832<br>(48,928,785; 70,504,878) |
| *Food Supplements* | 40,775,630 | 159,940 | 45,977,421<br>(32,065,523; 59,889,319) |

Notes: Scenarios highlighted with a light green background are non-dominated and lie on the cost-effectiveness frontier (Fig 4 in Results section). Values represent means across 30 stochastic runs; the 95% CIs reflect model stochasticity.

**Fig B4.**
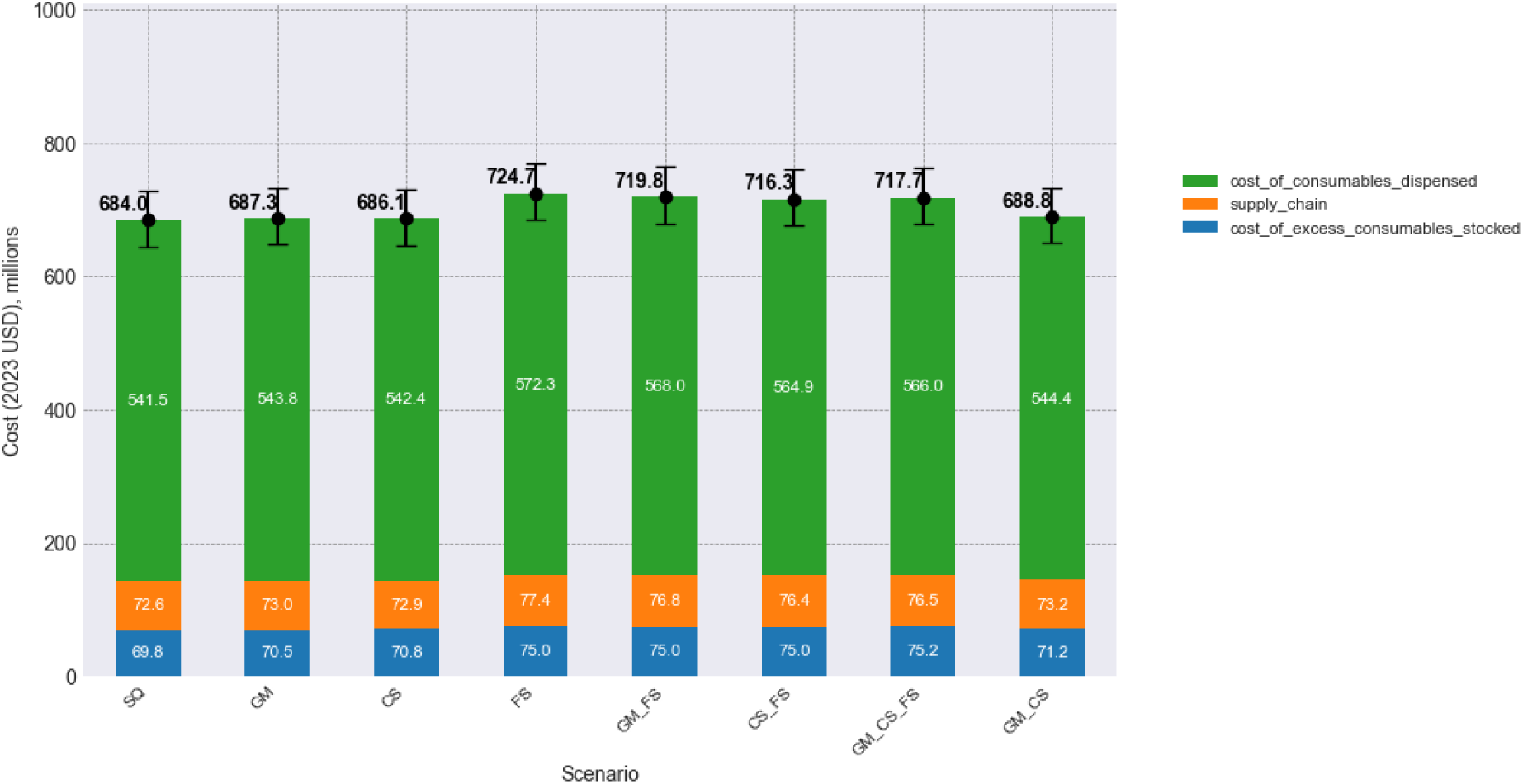
Consumable-related costs disaggregated by cost category.

The methodology for deriving these additional implementation costs is based on several simplifying, yet conservative, assumptions. We assume that the Growth Monitoring and Care-Seeking interventions target parents of newborns. For joint implementation, we assume a partial operational cost-sharing of 50%, reflecting delivery synergies; this is based on a meta-analysis showing that integrated disease management programmes can reduce costs to 75.9%, implying that 48.2% of implementation costs are shared [32]. A ‘no cost-sharing’ scenario is also explored in the sensitivity analysis below. Implementation costs for Food Supplements are assumed to scale linearly with the number of malnourished children.

We apply the full baseline unit cost for any intervention implemented in isolation. The unit cost multipliers for joint Growth Monitoring and Care-Seeking are: 1.0 (single intervention), 1.5 (joint with 50% cost-sharing), and 2.0 (joint with no cost-sharing).

For the Food Supplements intervention, a range of unit cost multipliers is explored in the sensitivity analysis to investigate the potential parameter space. This is necessary as specific implementation costs for Food Supplements are uncertain, given that the baseline unit costs are derived from education and promotion interventions.

Baseline costs were estimated from two primary sources. First, we adapted a unit cost of $3.56 (2014 USD) for promotion and education interventions in Bangladesh [33]. This was adjusted to $0.37 per parent per year (2023 USD) for the Malawian context, following the recommended approach to inflate the cost of non-tradable resources in low- and middle-income countries (LMICs) [34]. The unit cost was inflated via local currency units to 2023, converted back to USD, and translated between countries [35–37]. As the original estimate covered service delivery but excluded start-up costs, we assume the $0.37 covers 70% of first-year costs, with an additional 30% added for start-up (resulting in a $0.53 unit cost in the first year and $0.37 in subsequent years). Since the source estimate is for a one-year intervention, an annual discount rate of 3% is applied to subsequent years.

Second, we utilised a Malawi-specific estimate for joint promotion and education interventions [38]. The baseline estimate of $160 (2017 USD) is assuming wage rate of 50% of minimum wage, training every year, and calculated per preschool child targeted. However, if the training is assumed only in the first year of a five year long intervention period, the unit cost is reduced to $63, or if calculated per beneficiary (all children under five in intervention households, not only the primarily targeted preschool children), then to $41 [38]. Starting from a baseline of $160 (2017 USD), we adjusted the figure by assuming a wage rate of 50% of the minimum wage, training only in the first year, and costs per beneficiary (based on the sensitivity analysis within that study). To maintain the healthcare provider (MoH) perspective, we excluded volunteer time. Furthermore, meal provision was excluded as this is already covered by the *Wasting* model. Finally, we averaged the costs per intervention to allow for separate modelling. This results in a unit cost of $9.17 per parent of a newborn per year (2023 USD). As the original estimates incorporate start-up expenditures and 3% annual discounting, this value is applied consistently across all years. While we sought to exclude societal-perspective costs, some elements may remain captured within sub-cost categories; consequently, this figure serves as a conservative upper bound that likely exceeds the actual unit cost incurred by the MoH.

Total incremental costs and cost-effectiveness (using a CET of $76 per DALY averted) were analysed for the scenarios defined in <u>Table B4</u>. We incorporated sensitivity to: (i) the baseline unit cost estimate (using both estimates explained above); (ii) the assumption of shared implementation costs for joint Growth Monitoring and Care-Seeking (50% or 0%); and (iii) a range of unit cost multipliers (10, 2, 1, and 0.5) for the Food Supplements intervention.

Finally, the resulting annual costs were compared to the latest expenditure data for nutritional programmes in Malawi (FY 2018/19, inflated to 2023 USD [39]; <u>Table B7</u>) to identify which cost estimates reflect realistic national spending levels. The ranges of modelled total incremental costs, normalised per intervention included in a scenario, are provided in <u>Table B8</u>.

Expenditure data indicate that the mean annual spend on a nutritional programme was $636,178 (<u>Table B7</u>), consequently, the modelled total incremental costs using the lower baseline unit costs of $0.53 (first year) and $0.37 (subsequent years) [33] appear most realistic (<u>Table B8</u>). Furthermore, sensitivity analysis (<u>Fig B5</u>) demonstrates that results are qualitatively robust to variations in shared implementation costs and the Food Supplements multiplier, with the primary driver of outcome variance being the choice of baseline unit cost. Except, when higher baseline unit cost is applied, then multiplier of unit cost for Food Supplements determines whether no scenario, or Food Supplements scenario is cost-effective. Therefore, for the main cost-effectiveness analysis, we adopted the following assumptions: baseline unit cost of $0.53 in the first year and $0.37 in subsequent years [33]; 50% shared implementation costs for joint Growth Monitoring and Care-Seeking delivery; and the full baseline unit cost for Food Supplements implementation (FS multiplier of 1). The results of this main analysis are presented in the *Results* section and highlighted within the three-way sensitivity analysis (<u>Fig B5</u>).

**Table B7.** Expenditure per a nutritional project FY 2018/19 in Malawi.

| Statistic | Expenditure (2023 USD) |
| --- | --- |
| min | 81 |
| max | 18,077,007 |
| mean | 636,178 |
| lowest 5 | 81; 357; 540; 581; 1,622 |
| highest 5 | 630,861; 713,790; 1,236,062; 2,232,115; 18,077,007 |

Note: Expenditure on all nutritional projects in FY 2018/19 was \$27,991,823 (2023 USD).

**Table B8.** Modelled average annual total incremental costs range (min; max) over all scenarios, per one intervention implemented within the scenario (2023 USD).

| Baseline unit cost;<br>GM & CS shared implementation costs<br>proportion | FS multiplier |  |  |  |
| --- | --- | --- | --- | --- |
|  | 10 | 2 | 1 | 0.5 |
| \$0.53 (first year), \$0.37 (subsequent years) [33];<br>GM & CS: 0.5 | 699,223;<br>8,475,006 | 699,223;<br>8,219,102 | 699,223;<br>8,187,114 | 699,223;<br>8,171,120 |
| \$0.53 (first year), \$0.37 (subsequent years) [33];<br>GM & CS: 0 | 716,034;<br>8,475,006 | 716,034;<br>8,219,102 | 716,034;<br>8,187,114 | 716,034;<br>8,171,120 |
| \$9.17 (each year) [38];<br>GM & CS: 0.5 | 5,744,809;<br>15,559,375 | 5,744,809;<br>9,635,976 | 5,744,809;<br>8,895,551 | 5,744,809;<br>8,525,339 |
| \$9.17 (each year) [38];<br>GM & CS: 0 | 7,443,796;<br>15,559,375 | 7,315,248;<br>9,635,976 | 7,034,286;<br>8,895,551 | 6,885,737;<br>8,525,339 |

Note: Under the assumption of a \$9.17 baseline unit cost, the threshold for the Food Supplements scenario to remain cost-effective (i.e. ICER < CET of \$76) is a Food Supplements multiplier of 1.4. This corresponds to a unit cost of \$12.8 for the Food Supplements intervention, above which the scenario is no longer cost-effective.

**Fig B5.**
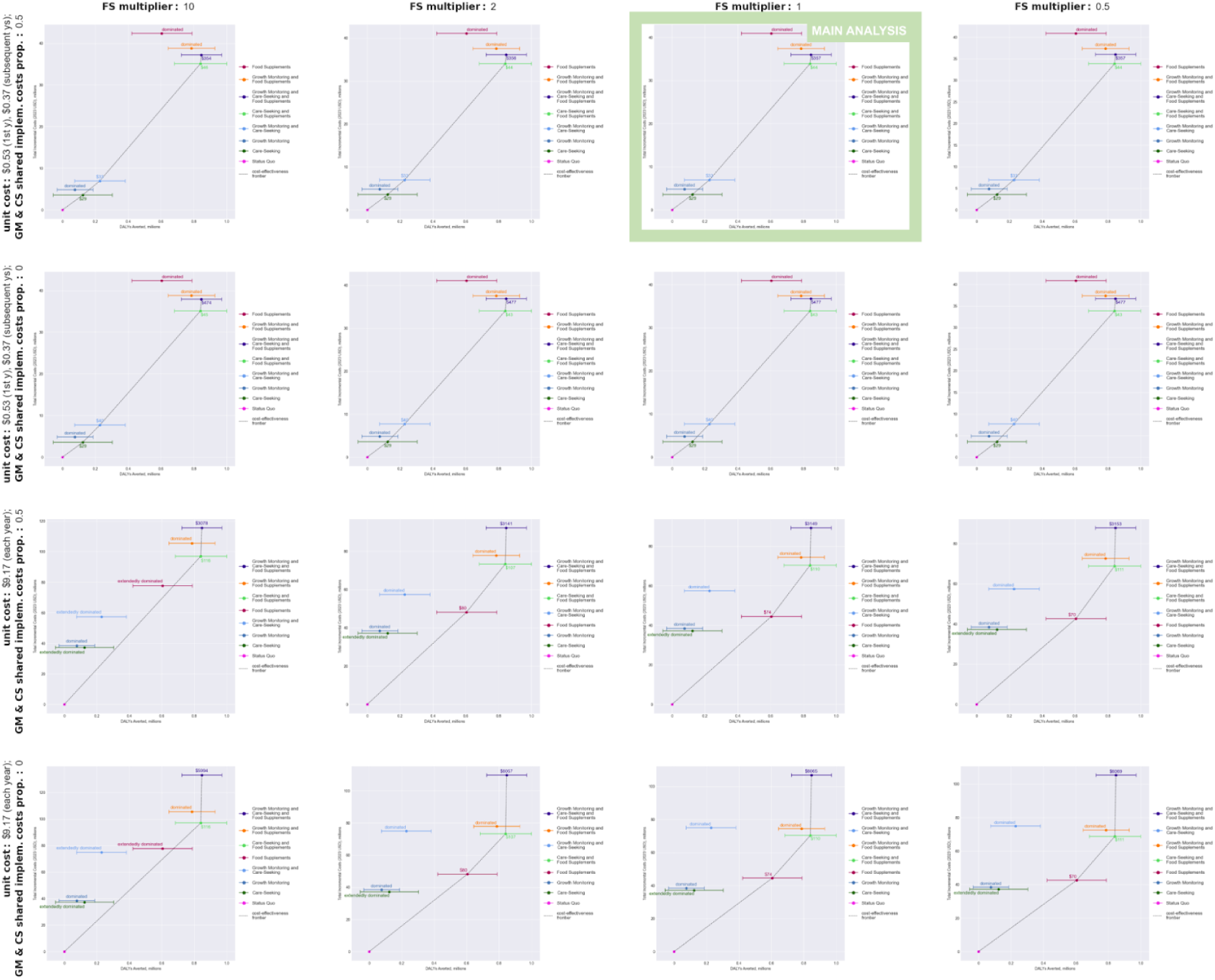
Cost-effectiveness plane: sensitivity to implementation costs. Note: Under the assumption of a $9.17 baseline unit cost, the threshold for the Food Supplements scenario to remain cost-effective (i.e. ICER < CET of $76) is a Food Supplements multiplier of 1.4. This corresponds to a unit cost of $12.8 for the Food Supplements intervention, above which the scenario is no longer cost-effective.

To assess the robustness of our main findings to additional implementation cost estimates, we calculated the maximum allowable total incremental costs at which a scenario remains cost-effective at a CET of $76 per DALY averted. By comparing this threshold to the total incremental costs estimates presented in the main analysis (Table 2 in *Results* section), we determined the ‘financial headroom’ or reserve for each scenario (<u>Table B6</u>). These provide a benchmark for policymakers regarding the feasibility of implementing the scenarios. For the optimal scenario (Care-Seeking and Food Supplements), our estimated total incremental costs of $33.9 million maintain a substantial reserve, as the strategy remains cost-effective up to $63.8 million (lower 95% CI bound: $51.8 million).

#### Appendix C

##### Moderate wasting incidence regression model

for those not having acute malnutrition, ie well-nourished, not treated at the moment, and not recovered at least 14 days before the incidence

1. Unscaled Wasting Incidence Model:

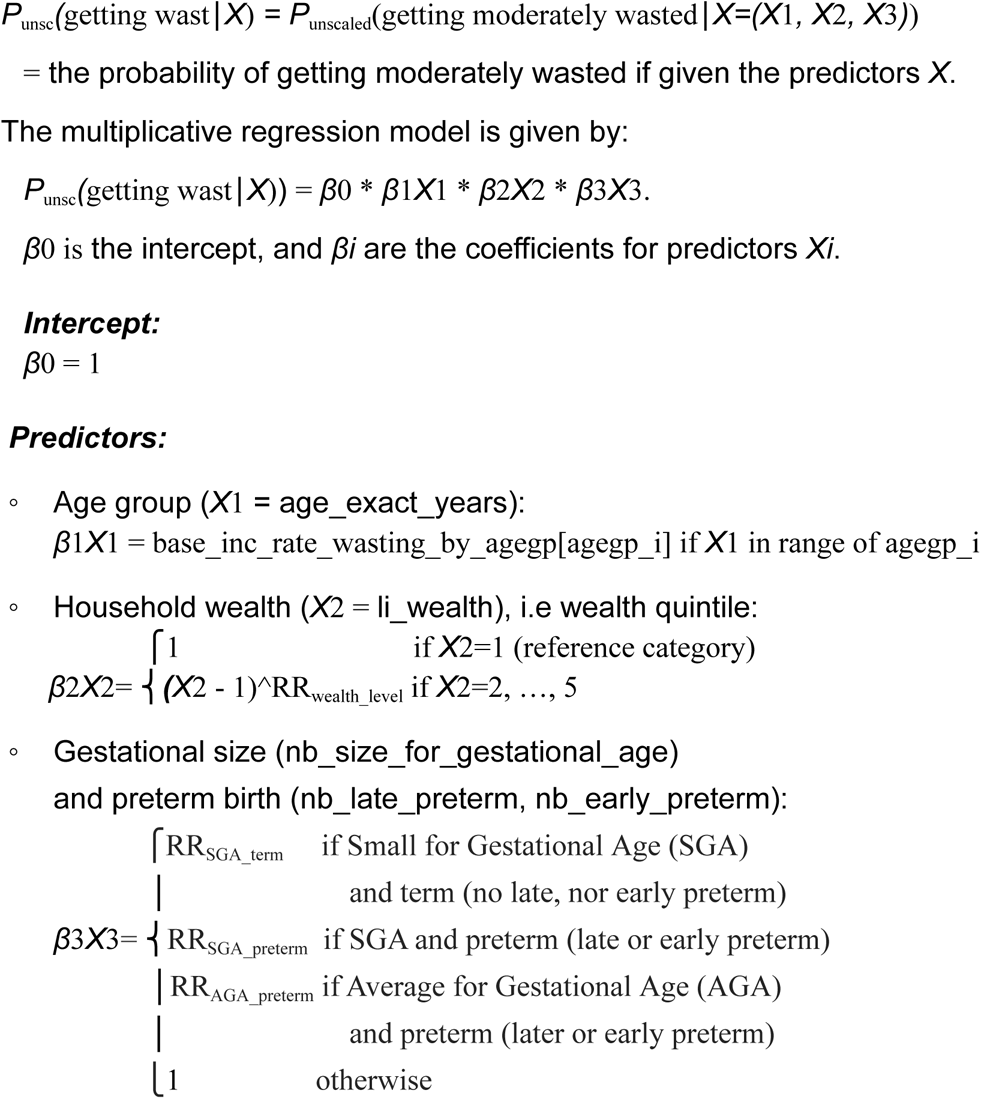
2. Intercept Scaling:

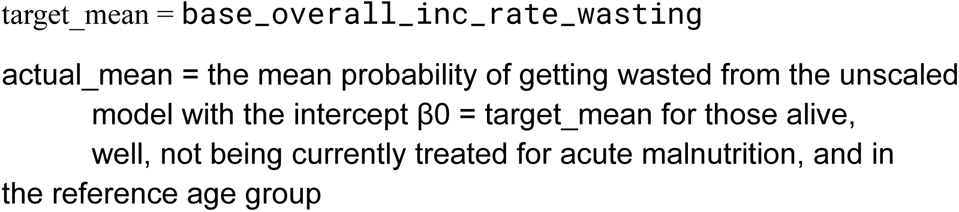

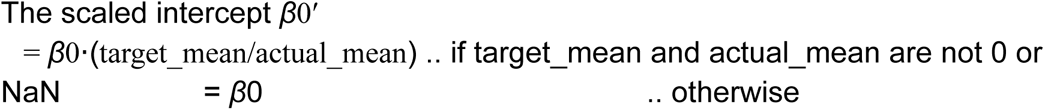
3. **Scaled Wasting Incidence Model**: The scaled multiplicative model is:

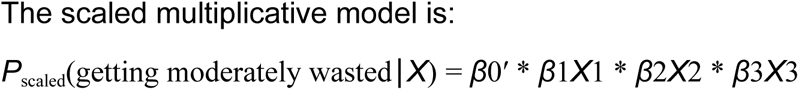

##### Age-specific relative risks of moderate wasting incidence and progression to severe wasting

We calibrated age-specific monthly risks for the onset of moderate wasting (MW) and the subsequent progression to severe wasting (SW) using a system of simplified discrete-time difference equations in order to derive age-specific relative risks *RR*_*MW,a*_ (rr_inc_rate_wasting_by_agegp) and *RR*_*SW,a*_ (rr_progression_rate_to_severe_wasting_monthly_by_agegp) to parameterise the *Wasting* model.

For each age group *a*, the number of wasted children at the month *t* + 1 is determined as follows:

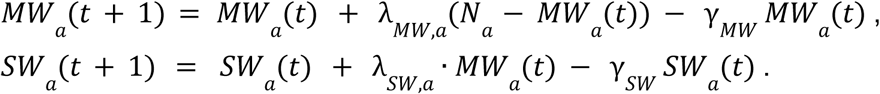

|  |  |
| --- | --- |
| $MW_a(t), SW_a(t)$ | the number of wasted children moderately or severely, respectively, in age group $a$, at time $t$ |
| $N_a(t)$ | the total number of children in age group $a$, at time $t$ |
| $\gamma_{MW}, \gamma_{SW}$ | the recovery rate of $1/D_{MW}$ or $1/D_{SW}$, respectively, where average duration $D_{MW}$ or $D_{SW}$ of moderate wasting or severe wasting, respectively, are common across all age groups |
| $\lambda_{MW,a}, \lambda_{SW,a}$ | the monthly incidence risks of moderate wasting onset or progression to severe wasting, respectively, both being calibrated |

The monthly risks (λ_*MW,a*_, λ_*SW,a*_), were calibrated to match the expected prevalence decrease after one month:

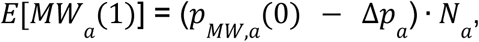

where Δ*p*_*a*_ is the observed average monthly decrease in prevalence for an age group *a,* derived from the trend between 2010 and 2019 [1,40].

The age-specific relative risks were then determined by comparing the calibrated risks to the reference category (the youngest age group):

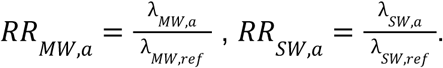

##### Modelled incidence cases of moderate and severe wasting

The following figure presents simulated annual incidence cases of moderate and severe wasting by age (0–4 years) from 2015 to 2030. These outputs were generated using the calibrated model described in the main text and reflect age-specific patterns over time under the status quo scenario. The figure illustrates both baseline trends and variability across age groups.

**Fig C1.**
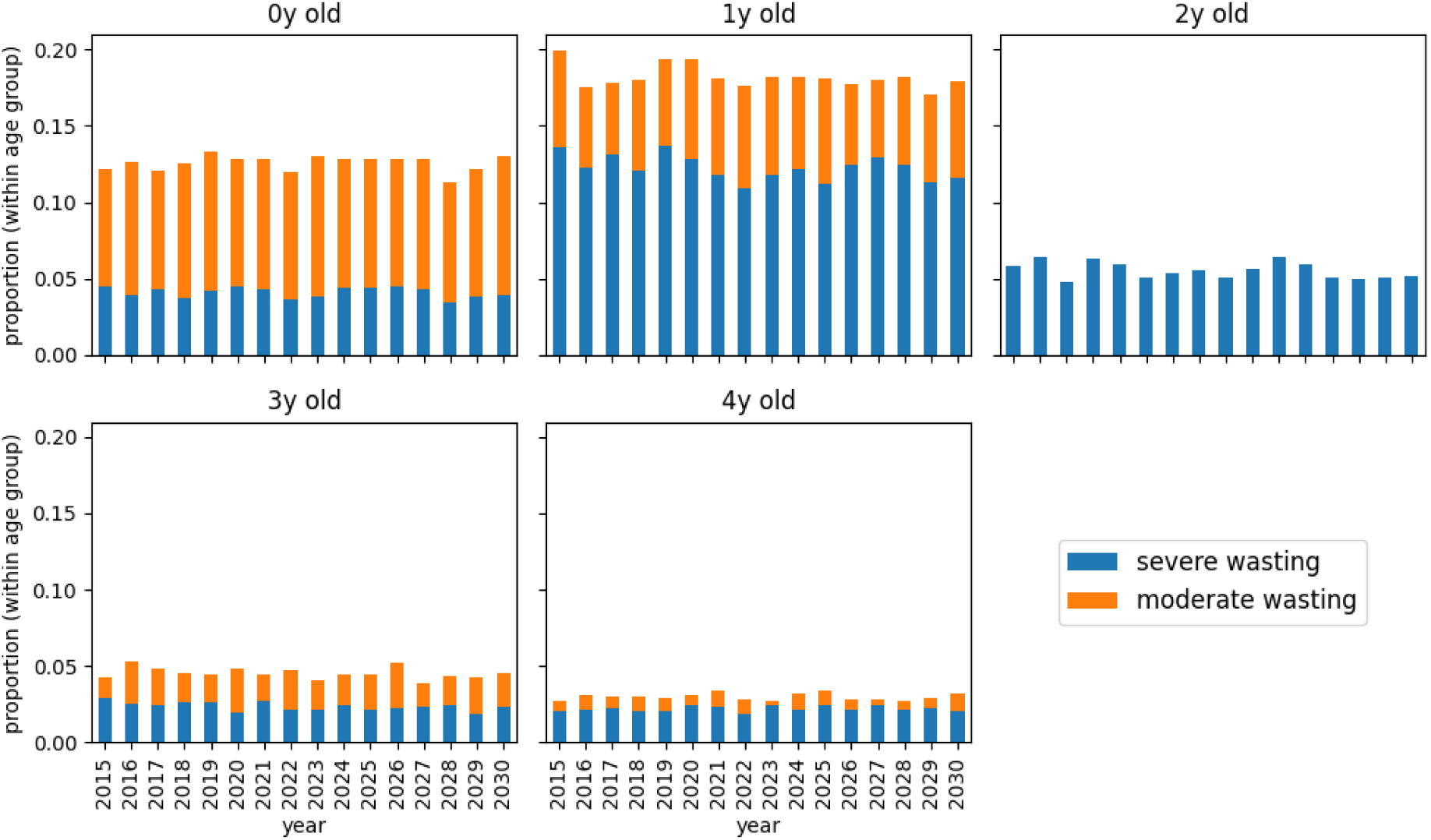
Annual incidence of wasting by age, 2015–2030.

#### Appendix D

##### Overall outcomes

**Fig D1.**
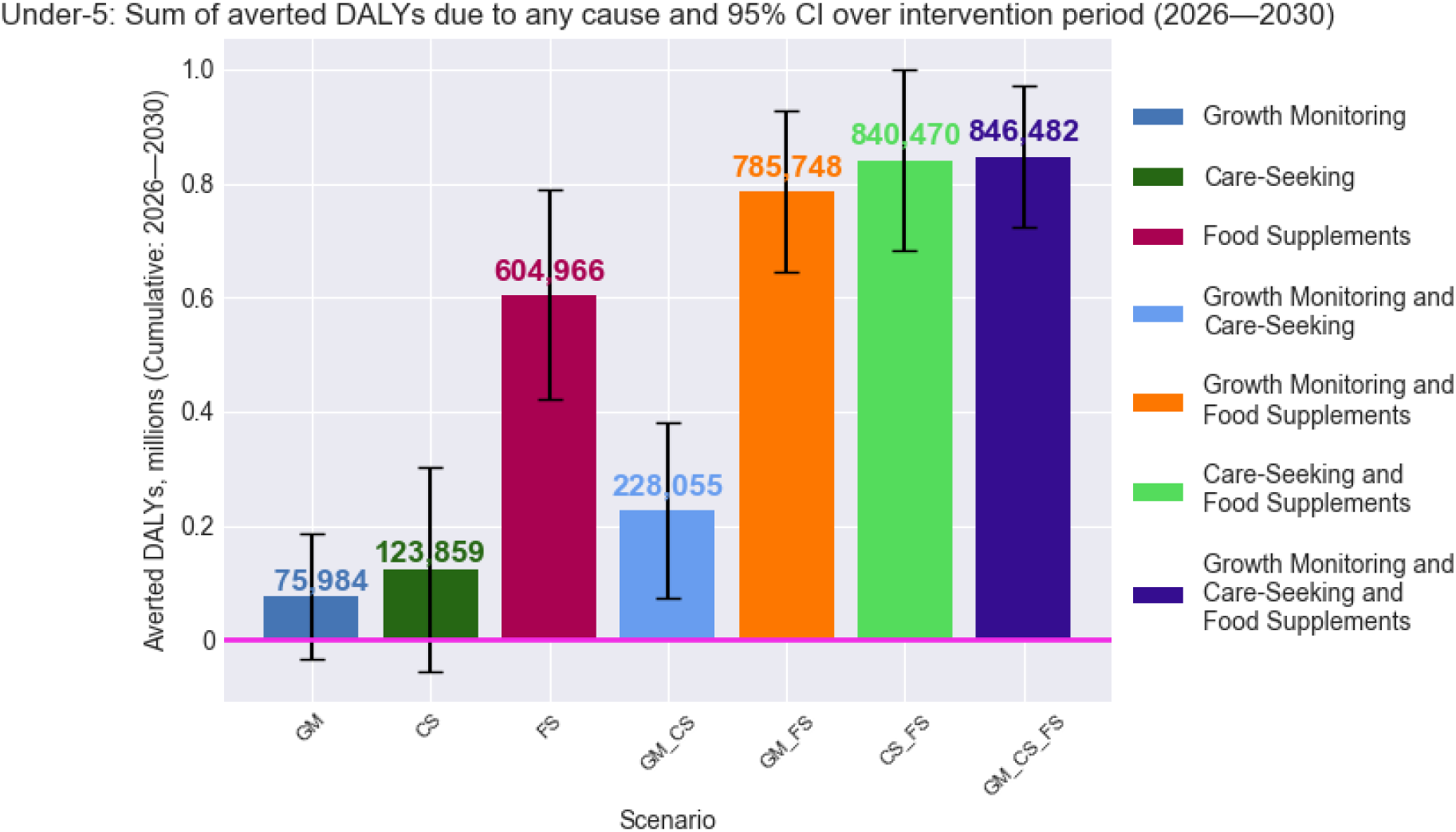
Overall averted DALYs by scenario (2026–2030).

##### Cause-specific outcomes

Cause-specific estimates show that the majority of averted DALYs across all intervention scenarios are attributable to reductions in severe acute malnutrition (SAM). As summarised in <u>Table D1</u>, all scenarios demonstrate significant reductions in SAM-related DALYs (<u>Fig D2</u>), underscoring the direct effectiveness of these interventions in addressing acute malnutrition, however, there is no observable effect on DALYs due to acute lower respiratory infections (ALRI) (<u>Fig D3</u>) or diarrhoea (<u>Fig</u> <u>D5</u>).

Similar outcomes are observed regarding averted deaths (now shown), however when disaggregated to ALRI- and diarrhoea-related deaths among children with SAM (<u>Fig D4</u> and <u>Fig D6</u>, respectively), more consistent and statistically significant reductions are observed. These findings suggest that the interventions have an effect on the ALRI- and diarrhoea deaths, however, their overall impact remains limited.

This limited indirect effect is likely explained by the low prevalence of SAM among children dying from ALRI and diarrhoea under the status quo, only 3.8% and 5.4%, respectively (not shown). This suggests that while SAM increases vulnerability to other illnesses, it is not the predominant underlying condition in most ALRI- or diarrhoea-related deaths.

**Table D1.** Cause-specific averted DALYs by scenario (2026–2030).

| Scenario | DALYs averted vs SQ (95% CI)<br>by cause associated with acute malnutrition |  |  |
| --- | --- | --- | --- |
|  | SAM | ALRI | Diarrhoea |
| Growth Monitoring | 145,872<br>(105,076; 186,669) | 9,506<br>(-74,101; 93,114) | 4,920<br>(-39,938; 49,778) |
| Care-Seeking | 241,250<br>(183,302; 299,199) | -35,128<br>(-132,776; 62,520) | 4,894<br>(-48,341; 58,130) |
| Food Supplements | 704,262<br>(662,144; 746,380) | 72,176<br>(-34,716; 179,069) | 4,874<br>(-41,711; 51,459) |
| Growth Monitoring and Care-Seeking | 314,068<br>(253,857; 374,280) | 12,734<br>(-67,056; 92,525) | -6,417<br>(-65,736; 52,903) |
| Growth Monitoring and Food Supplements | 823,126<br>(781,659; 864,594) | 13,791<br>(-66,960; 94,541) | 30,241<br>(-20,025; 80,506) |
| Care-Seeking and Food Supplements | 800,263<br>(758,746; 841,780) | 12,612<br>(-68,492; 93,715) | 37,633<br>(-6,082; 81,349) |
| Growth Monitoring and Care-Seeking and Food Supplements | 837,458<br>(797,550; 877,366) | 74,297<br>(-21,021; 169,615) | 34,908<br>(-17,293; 87,109) |

###### SAM-related outcomes

**Fig D2.**
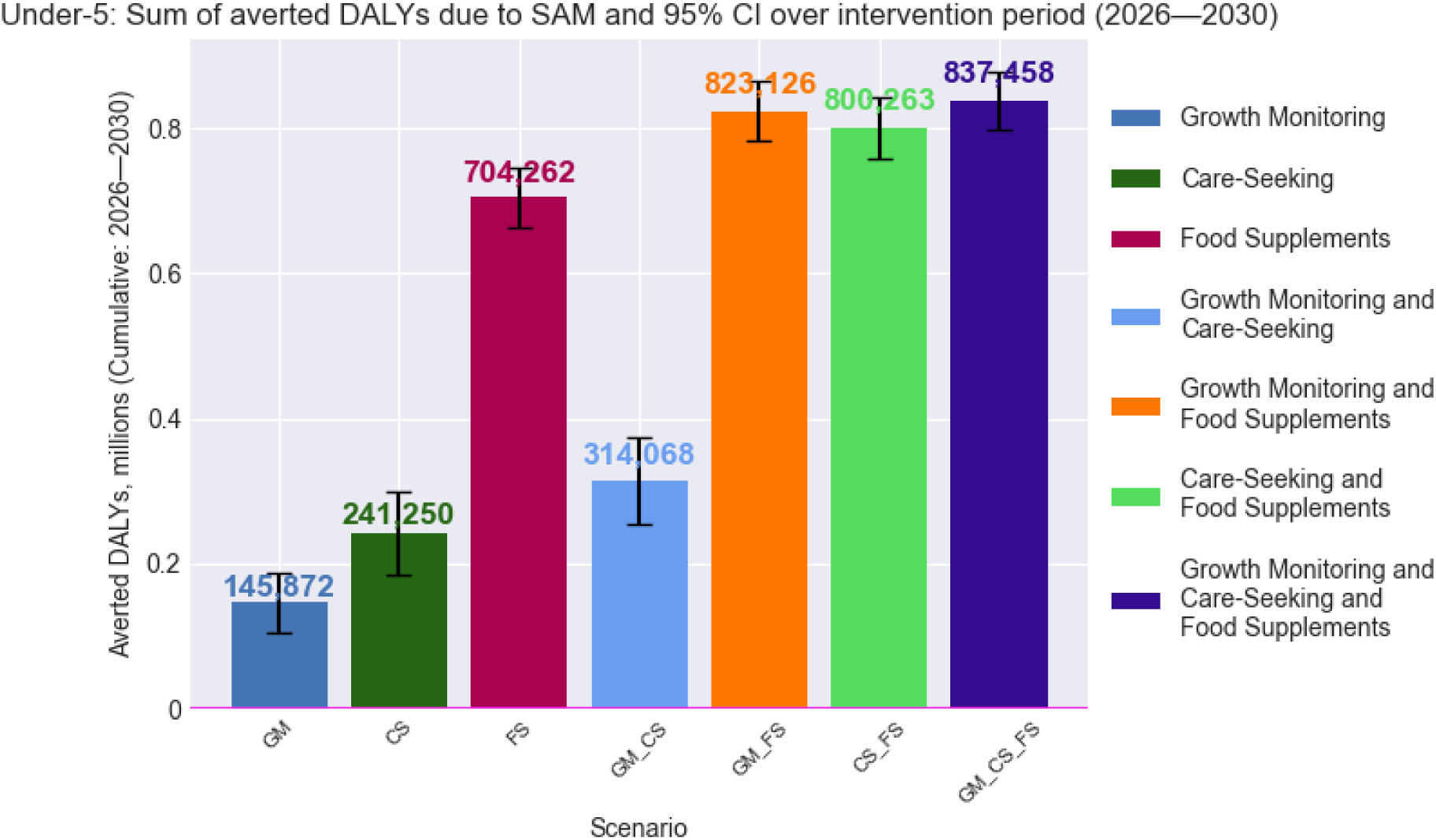
Averted DALYs due to SAM.

###### ALRI-related outcomes

**Fig D3.**
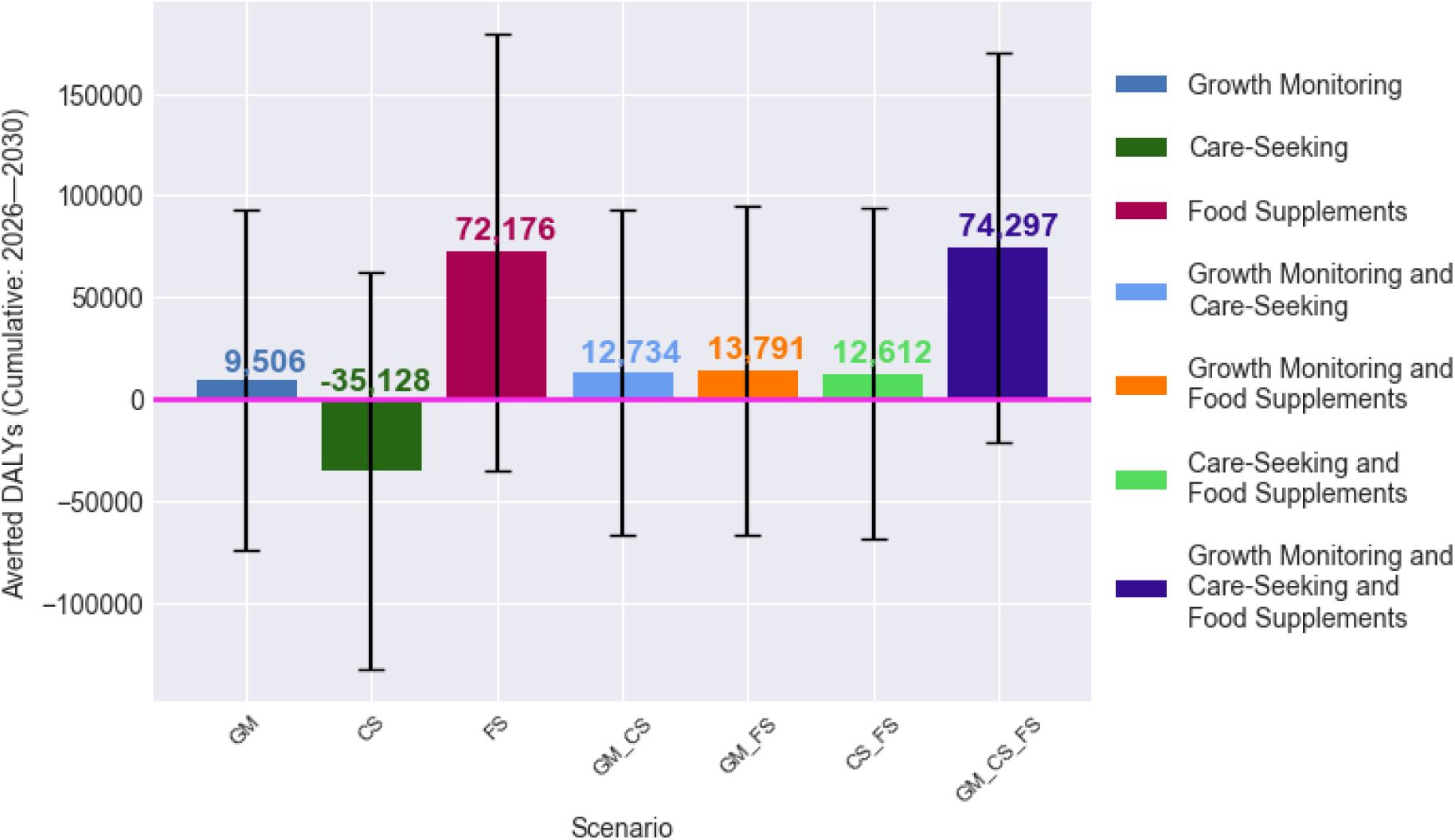
Averted DALYs due to ALRI.

**Fig D4.**
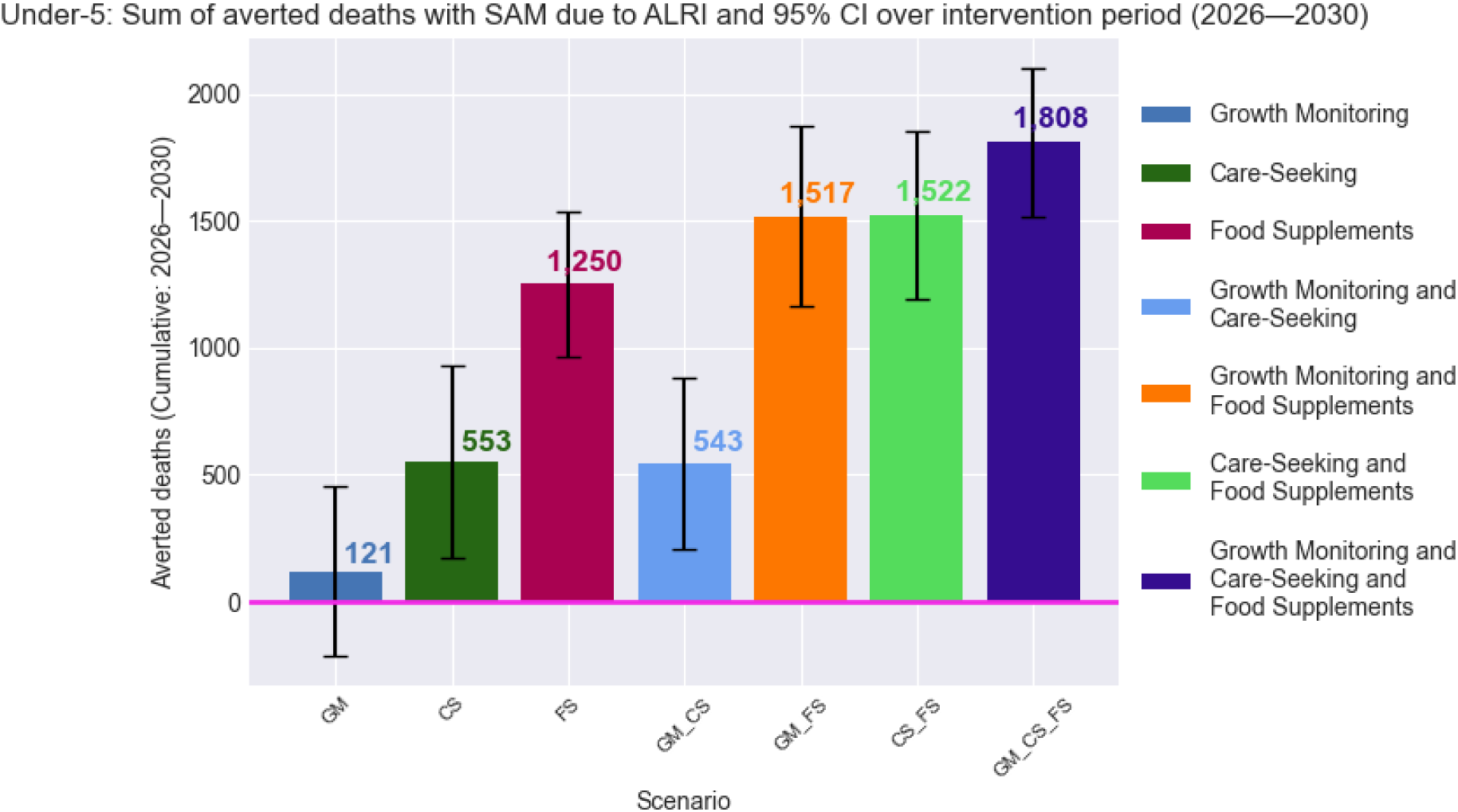
Averted deaths due to ALRI in children with SAM.

###### Diarrhoea-related outcomes

**Fig D5.**
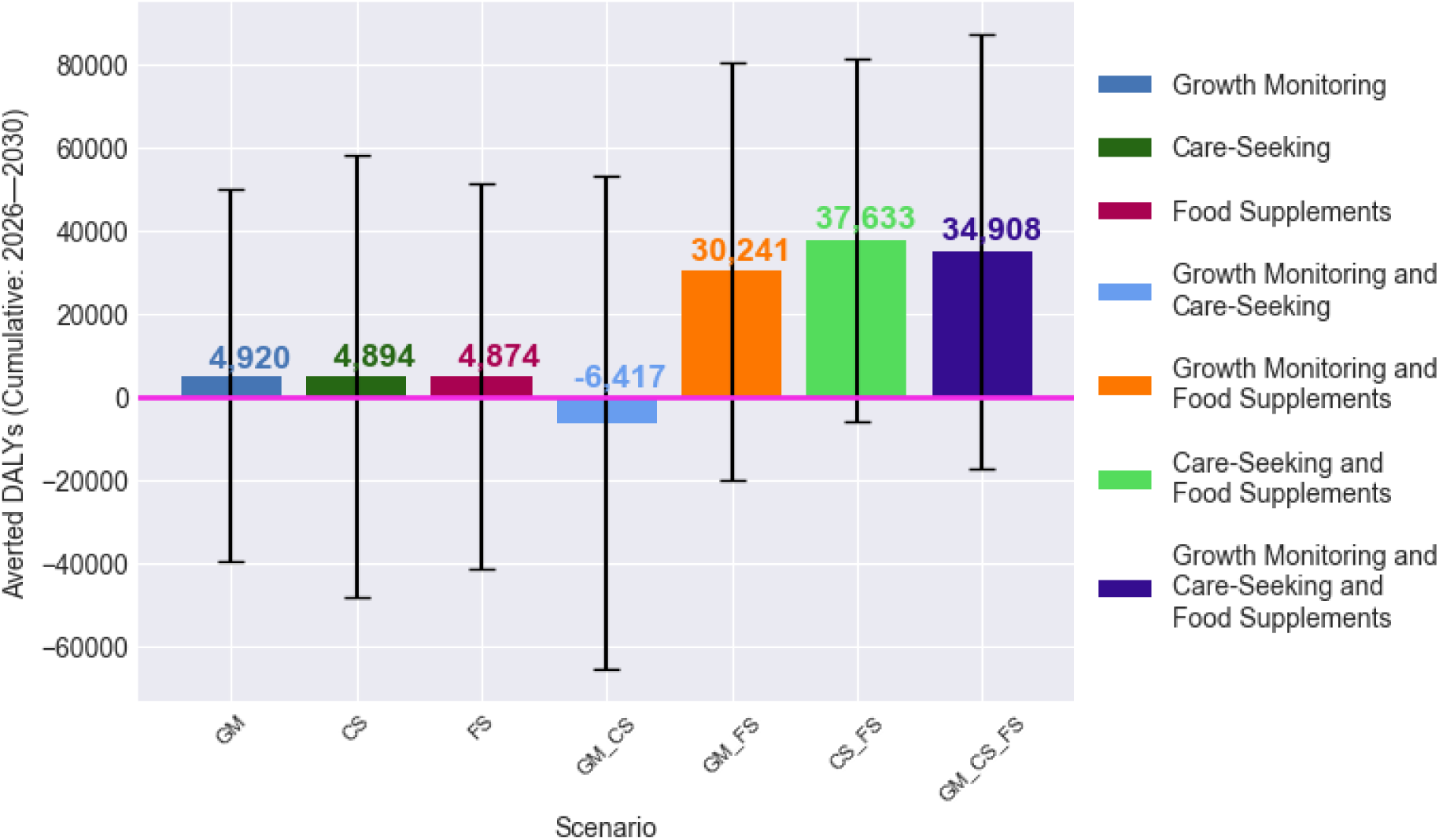
Averted DALYs due to diarrhoea.

**Fig D6.**
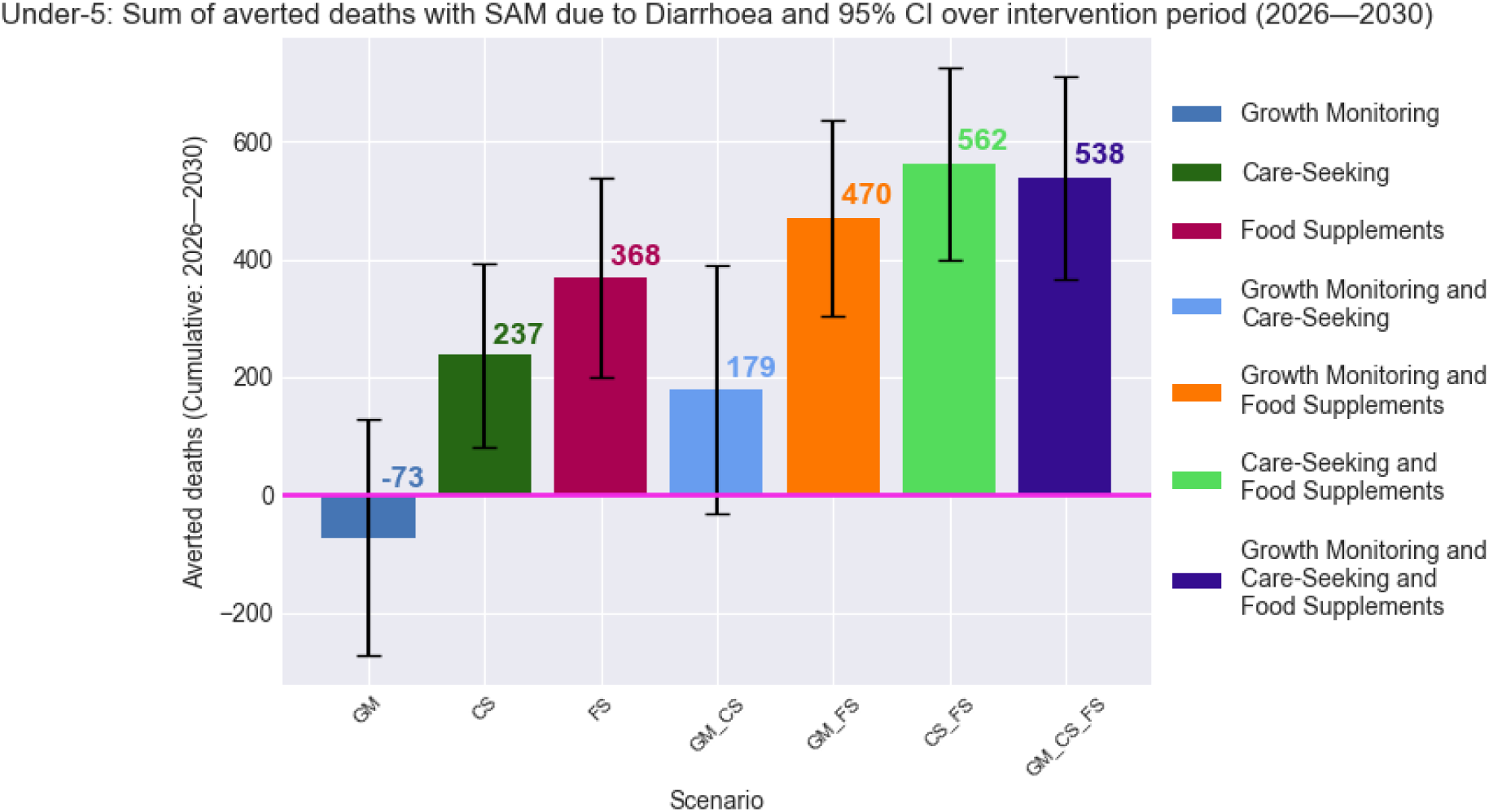
Averted deaths due to diarrhoea in children with SAM.

